# Urinary high-risk HPV DNA detection to enhance cervical cancer screening in developing countries

**DOI:** 10.1101/2023.10.26.23297586

**Authors:** Novia Syari Intan, Revata Utama, Dewi Wulandari, Reiva Wisdharilla, Shafira Mutia Khanza, Muhamad Rifki Ramadhan, Indah Suci Widyahening, Neni Nurainy, Rini Mulia Sari, Andrijono

## Abstract

**Objectives:** To increase cervical cancer screening capacity and participation, we evaluated the performance of the newly developed hrHPV ReadyMix qPCR Kit for detecting high-risk Human Papillomavirus (HPV) in urine samples while simultaneously genotyping HPV16, HPV18, and HPV52.

**Methods:** 876 samples were used to assess the performance of hrHPV ReadyMix qPCR Kit in detecting high-risk HPV in standard cervical swab sample compared to the Roche cobas^®^ 6800 HPV. The high-risk HPV detection in urine was compared to the corresponding paired cervical swab.

**Results:** The sensitivity of HPV detection in cervical swabs using hrHPV ReadyMix qPCR Kit reached 96.55% and the specificity reached 99.87%. Despite higher Ct values, urine samples demonstrated 80.88% sensitivity and 100.00% specificity compared to cervical swabs. Our method enables population-based high-risk HPV analysis with a 6.62% HPV prevalence from cervical swabs and 6.28% from urine samples. Furthermore, urine samples using the hrHPV ReadyMix qPCR Kit showed comparable HPV type distribution and the ability to genotype HPV16 and HPV18, to Roche the cobas® 6800 HPV.

**Conclusions:** Self-collected urine samples offer a 98.48% diagnostic accuracy for detecting high-risk HPV infection. This study highlights the hrHPV ReadyMix qPCR Kit’s potential in enhancing cervical cancer screening, offering valuable insights for future interventions.

## INTRODUCTION

Cervical cancer is the fourth most prevalent type of cancer among women, with approximately 604,127 cases and 341,831 fatalities worldwide in 2020.^1^ The distribution of cervical cancer cases varies across the globe, with developing countries accounting for 85% of the mortality^2^. In Indonesia, it is estimated that there are approximately 36,633 cases and 21,003 deaths related to cervical cancer per year.^3^ However, the disease is preventable and treatable if detected early. As over 99% of precancerous lesions (cervical dysplasia) and cervical carcinomas are caused by high-risk HPV infection,^4^ detection of high-risk HPV as the etiologic factor of cervical cancer cases has emerged as a primary early-screening method due to its greater sensitivity than the Pap test for detecting precancerous lesions of the cervix.^5,6^ It also offers the advantage of extended screening intervals from once every 3 years to once every 5 years as well as eliminating the need for skilled cytopathologists to interpret the results^4,5^.

However, implementation of HPV-based diagnostics in emerging countries faces a multitude of challenges, such as the high costs of screening tests, shortages of the appropriate diagnostic kits, and a lower than 10% screening participation rate in such countries, such as Myanmar and Indonesia.^7,8^ To increase the low participation rates, a self-sampling approach combining detection of HPV from urine samples with the utilization of the currently available infrastructure of quantitative polymerase chain reaction (qPCR) instruments was developed as an alternative to cervical swab-based HPV screening method. The use of urine samples offers several advantages, such as more privacy and less embarrassment in the sample collection process, reduced discomfort and pain, as well as ease of use. Furthermore, previous studies have shown that urine samples have a promising accuracy for HPV detection.^9–11^ To address those needs, the hrHPV ReadyMix qPCR Kit (ReadyMix) has been developed to detect 14 high-risk HPV types with the capability to genotype HPV52, HPV16, and HPV18 in both cervical swabs and urine samples. HPV16 and HPV18 contribute to about 70% of all cervical cancer cases. Alongside HPV52, they constituted the highest prevalence in Southeast Asia countries.^4,12^ Our study aimed to evaluate the performance of HPV detection in urine samples as compared to standard cervical swabs using the ReadyMix, which was assessed for performance against the Roche cobas^®^ 6800 HPV system. The insights gained from this study will contribute to the effort to achieve effective screening measures, not only in Indonesia but also in other countries with similar settings, thus reducing the burden of the disease.

## MATERIALS AND METHODS

### Ethics approval and consent to participate

The collection of clinical samples, cervical swabs and urine samples, was approved by the Health Research Ethics Committee of the Faculty of Medicine Universitas Indonesia Dr. Cipto Mangunkusumo Hospital under the identifier No. KET- 674 / UN2.F1/ETIK/PPM.00.02/2022. Informed consent was obtained from all participants and all methods were performed in accordance with the Declaration of Helsinki and the approved Indonesian guidelines and regulations for biomedical research.

### Study Design and Population

This cross-sectional diagnostic test accuracy study was conducted based on HPV detection results from a study population comprising sexually active women aged 20 to 50 years residing in three major Indonesian cities: Jakarta, Bandung, and Semarang. The minimum number of participants was determined based on an anticipated diagnostic sensitivity of 90% and a 4.4% hrHPV prevalence,^13^ yielding a requirement for a minimum of 786 participants.^14^ The study focused on women who sought healthcare services for general medical check-ups, Pap smears, or VIA test and agreed to take part in the study. The subjects were asked to provide urine and cervical swab samples. Pregnant women, those currently menstruating, and individuals who had already received the HPV vaccine were excluded from the study. This recruitment was conducted between July 2022 and September 2022.

### Specimen Collection

The participants were first instructed to collect their first-void urine, collected at any time of the day, in a urine container. The cervical swab was then taken with a physician’s assistance using a cervical brush and preserved in PreservCyt ® Solution (Hologic, Belgium). Both samples were kept at a temperature between 2-8°C and sent to the Clinical Pathology Department laboratory at the Cipto Mangunkusumo National Hospital for further analysis.

### DNA Extraction

The DNA was extracted from both urine and cervical swab samples using Zybio Nucleic Acid Extraction Kit (Zybio Inc., China) as per the manufacturer’s instructions for ReadyMix. The integrity of the extracted DNA was assessed by detecting the β-globin gene, later in the qPCR stage. The samples were simultaneously run in cobas®6800 HPV (Roche, Switzerland) where extraction and amplification are integrated in automatic system.

### HPV qPCR and Genotyping Protocol

The purified DNA was subjected to qPCR assays using ReadyMix performed in the CFX96™ System (Bio-Rad Lab, CA, USA) and using cobas^®^ 6800 HPV (Roche, Switzerland) (cobas) directly in the system. ReadyMix was developed by Nusantics and commercialized under the name of PathoScan hrHPV qPCR Kit (Nusantics, Indonesia) and CerviScan HPV-hr qPCR (PT Biofarma, Indonesia). hrHPV ReadyMix qPCR Kit targets the E6-E7 region to identify 14 high-risk HPV types while concurrently differentiating between HPV16, HPV18, and HPV52. Other HPV types targeted include HPV 31, 33, 35, 39, 45, 51, 56, 58, 59, 66, and 68, collectively detected in one fluorescence channel. Both tests detect human β-globin gene as an internal control (IC) to monitor the success of the extraction and qPCR procedure. cobas assay was performed according to the manufacturer’s instructions. ReadyMix reagent was pre-mixed in a single tube and thus can be directly aliquoted to qPCR well in a volume of 15 µL for a 20 µL qPCR reaction. This kit was then also used as per manufacturer recommendation, with PCR thermal cycles consisting of 95°C initial denaturation for 2 minutes and 45 cycles of 1-second denaturation at 95°C and 10 seconds annealing/extension at 60°C. The cutoff Ct value for a sample to be considered positive is 40.

In samples that show discordant results between the two testing methods or between different sample types, the presence of HPV DNA is confirmed using the amplicon sequencing method utilizing NGS technology. DNA libraries were amplified by PCR, prepared using Nextera XT index kit v2 (Illumina, CA, USA), and purified using AmPure XP beads (Beckman-Coulter, CA, USA). Paired-end (2x150 bp) sequencing was performed using the Illumina Miseq Micro v2 reagent kit (Illumina, CA, USA) on the MiSeq platform. The sequencing primers used have been previously validated through alignment between the sequencing results of synthetic genes and their corresponding references. Reads were demultiplexed based on their index, resulting in a pair of forward and reverse fastq file for each sample. The downstream data analysis was performed using QIIME2 platform.^15^ Primer sequences were trimmed and reads were filtered based on their length using cutadapt,^16^ with a cutoff length of 20. Forward and reverse reads were merged using vsearch,^17^ allowing a maximum of 5 mismatches. In this step, a minimum merged read length of 30 for E6-E7 was enforced. Merged reads were quality-filtered based on their Q score with a cutoff value of 30 prior to dereplication. Reads were clustered *de novo* to produce operational taxonomic units with 99% similarity. Chimera sequences were filtered out using vsearch utility uchime-denovo. The resulting sequences were classified into either of HPV types by mapping them to HPV reference sequences that are obtained from Papillomavirus Episteme (PaVE) (https://pave.niaid.nih.gov/). The mapping was performed using BLASTn with a minimum of 90% identity and a minimum word size of 20. ^18^

### Statistical analysis

The data were assessed for normality using the Shapiro-Wilk test. Categorical variables were presented as percentages. For measures of central tendency, the mean ± standard deviation (SD) was applied to normally distributed data, while the median (interquartile range; IQR) was employed for non-normally distributed data. The study calculated the sensitivity, specificity, positive predictive value (PPV), negative predictive value (NPV), and diagnostic accuracy to evaluate the diagnostic performance. Cohen’s Kappa coefficient (κ) was used to determine concordance between compared tests or samples.^19^ The Wilcoxon matched-pairs signed rank test was performed to compare qPCR Ct values between urine and cervical swabs. All statistical analyses were executed using GraphPad statistical software version 10.0.2 (GraphPad San Diego, CA, USA).

## RESULTS

### Summary of subject participation and data validity

Between July and October 2022, 901 women who visited several healthcare facilities in Jakarta, Bandung, and Semarang for general medical check-ups, Pap smears, or visual inspection with acetic acid (VIA) tests and expressed interest in participating in this study were evaluated for eligibility. Sixteen individuals aged older than 50 years were consequently excluded. Among the remaining 885 participants engaged in sample collection, nine were ineligible due to incomplete collection of paired cervical swabs and urine samples. Therefore, a total of 876 subjects were included in this study (Fig. 1), consists of 157 subjects within the age group 20-29 (17.92%), 346 subjects within the age group 30-39 (39.50%), and 373 subjects within the age group 40-50 (42.58%). The mean age of the subjects was 37.23±7.28 years. As many as 385 subjects were from Jakarta (43.95%), 355 subjects from Bandung (40.53%), and 136 subjects from Semarang (15.53%).

**Figure 1.**
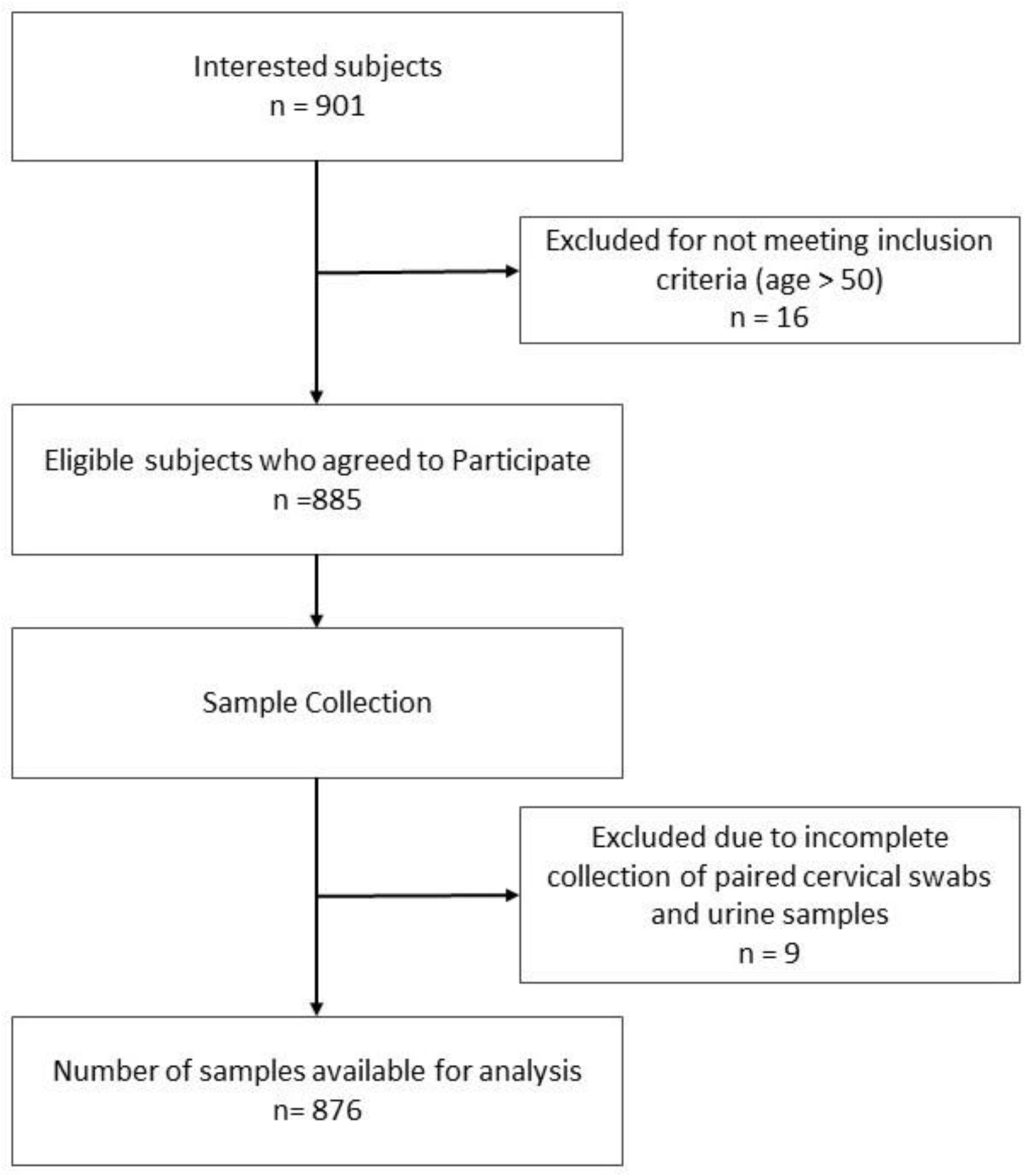
Flow chart describing study subjects’ participations

Among the 876 cervical swab and urine samples tested with cobas, there were 24 cervical swab samples (2.74%) and 206 urine samples (23.52%) that yielded invalid results (Table 1). On the other hand, ReadyMix produced a smaller number of invalid results, with only 2 cervical swab samples (0.23%) and 19 urine samples (2.17%) being invalid (Table 1).

**Table 1.** Summary of subject participation and data validity.

| Results | ReadyMix |  | cobas |  |
| --- | --- | --- | --- | --- |
|  | Cervical Swab | Urine | Cervical Swab | Urine |
| Total subjects | 876 | 876 | 876 | 876 |
| Subject with complete test result | 874 | 857 | 852 | 670 |
| Invalid | 2 | 19 | 24 | 206 |
| Invalid Rate | 0.23% | 2.17% | 2.74% | 23.52% |

### hrHPV ReadyMix qPCR Kit demonstrates high sensitivity and specificity in high-risk HPV detection

HPV detection in this study specifically refers to high-risk HPV detection. ReadyMix’s technical performance has been previously characterized based on linearity assay and limit of detection (LoD) which yielded satisfactory results (Supplementary Materials: Supplementary Fig. S1, S2). We first sought to determine the diagnostic performance of ReadyMix in HPV detection using the cervical swab as a standard method. The accuracy was excellent with a sensitivity of 96.15% (95% CI: 87.02-99.32%) and a specificity of 99.13% (95% CI: 98.2-99.58) (Table 2).

**Table 2.**
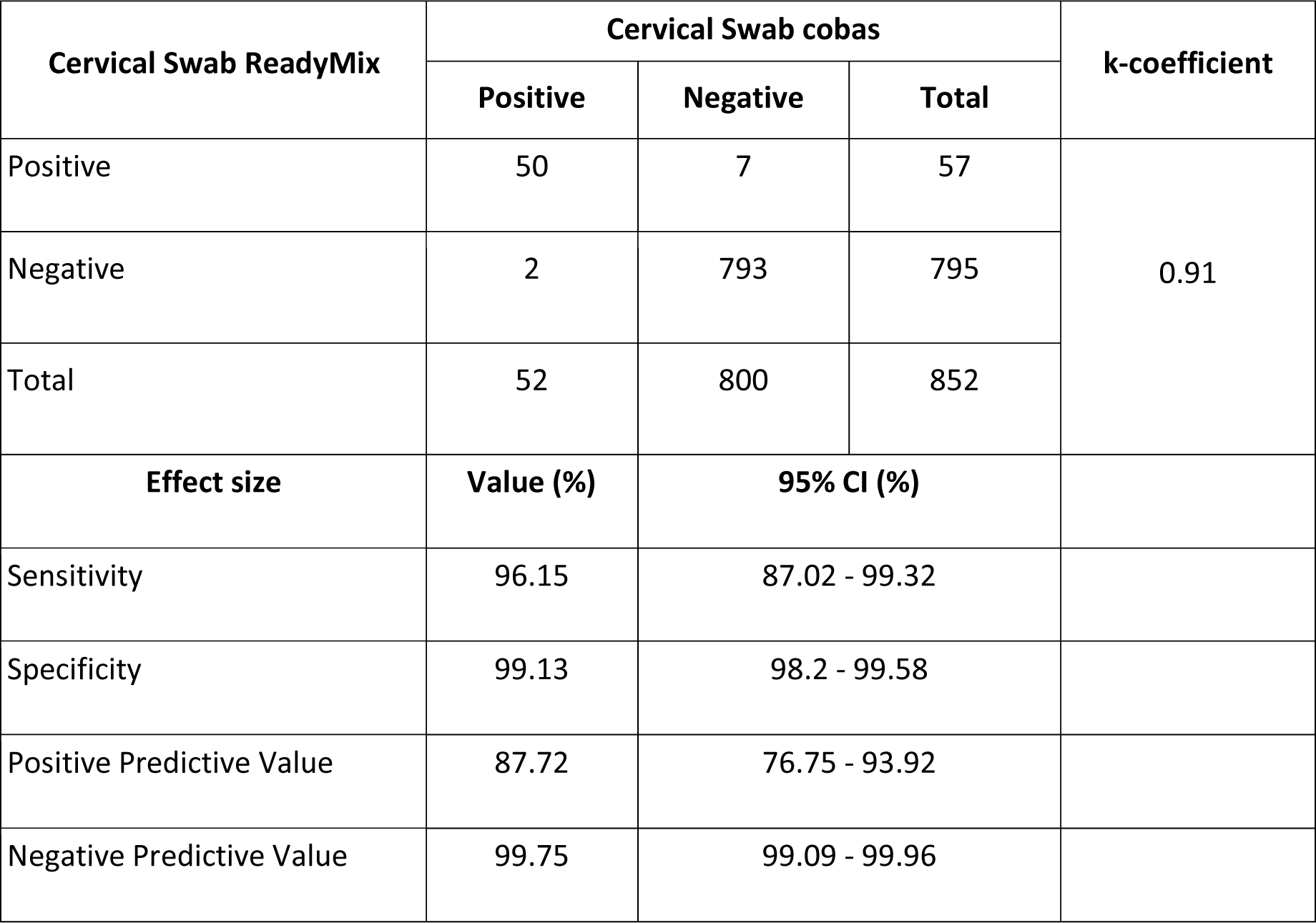
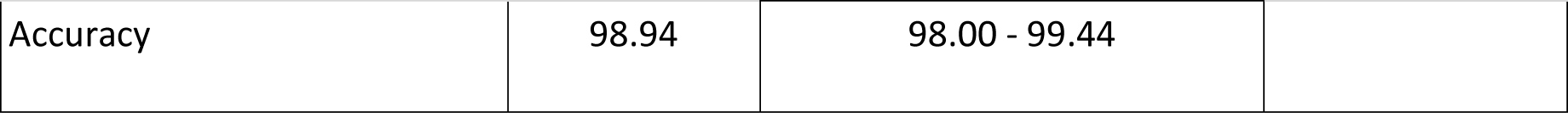
HPV detection in the cervical swab using ReadyMix versus cobas.

| Cervical Swab ReadyMix | Cervical Swab cobas |  |  | k-coefficient |
| --- | --- | --- | --- | --- |
|  | Positive | Negative | Total |  |
| Positive | 50 | 7 | 57 | 0.91 |
| Negative | 2 | 793 | 795 |  |
| Total | 52 | 800 | 852 |  |
| Effect size | Value (%) | 95% CI (%) |  |  |
| Sensitivity | 96.15 | 87.02 - 99.32 |  |  |
| Specificity | 99.13 | 98.2 - 99.58 |  |  |
| Positive Predictive Value | 87.72 | 76.75 - 93.92 |  |  |
| Negative Predictive Value | 99.75 | 99.09 - 99.96 |  |  |

|  |  |  |
| --- | --- | --- |
| Accuracy | 98.94 | 98.00 - 99.44 |

Upon observing samples with discordant results, we found that 6 out of 7 false-positives results were confirmed true-positives by the NGS results (Supplementary Materials: Supplementary Table S1). To obtain a clearer performance report of this assay, we then incorporate NGS data, which resulted in 96.55% (95% CI: 88.27-99.39%) sensitivity and 99.87% (95% CI: 99.29-99.99%) specificity (Table 3). Cohen’s Kappa analysis (0.97) indicated an almost perfect agreement between the compared methods.

**Table 3.** HPV detection in the cervical swab using ReadyMix versus cobas and NGS.

| Cervical Swab ReadyMix | Cervical Swab cobas & NGS |  |  | k-coefficient |
| --- | --- | --- | --- | --- |
|  | Positive | Negative | Total |  |
| Positive | 56 | 1 | 57 | 0.97 |
| Negative | 2 | 793 | 795 |  |
| Total | 58 | 794 | 852 |  |
| Effect size | Value (%) | 95% CI (%) |  |  |
| Sensitivity | 96.55 | 88.27 - 99.39 |  |  |
| Specificity | 99.87 | 99.29 - 99.99 |  |  |
| Positive Predictive Value | 98.25 | 90.71 - 99.91 |  |  |
| Negative Predictive Value | 99.75 | 99.09 - 99.96 |  |  |
| Accuracy | 99.65 | 98.97 - 99.90 |  |  |

### Urine samples exhibit an excellent agreement with cervical swabs in HPV detection

To validate the use of urine sample for HPV detection, we compared its performance with a well-established cervical swab sample. The NGS-corrected data were used to analyse the ReadyMix’s performance. The NGS results were attached in supplementary data (Supplementary Materials: Supplementary Table S1; Supplementary Fig.S3). We found an almost perfect agreement (κ: 0.89) between cervical swabs and urine with a sensitivity of 80.88% (95% CI: 69.99-88.47%) and 100% (95% CI: 99.51-100%) specificity (Table 4). This result demonstrates enhanced accuracy in urine HPV detection on ReadyMix as compared to cobas (Supplementary Materials: Supplementary Table S2).

**Table 4.**
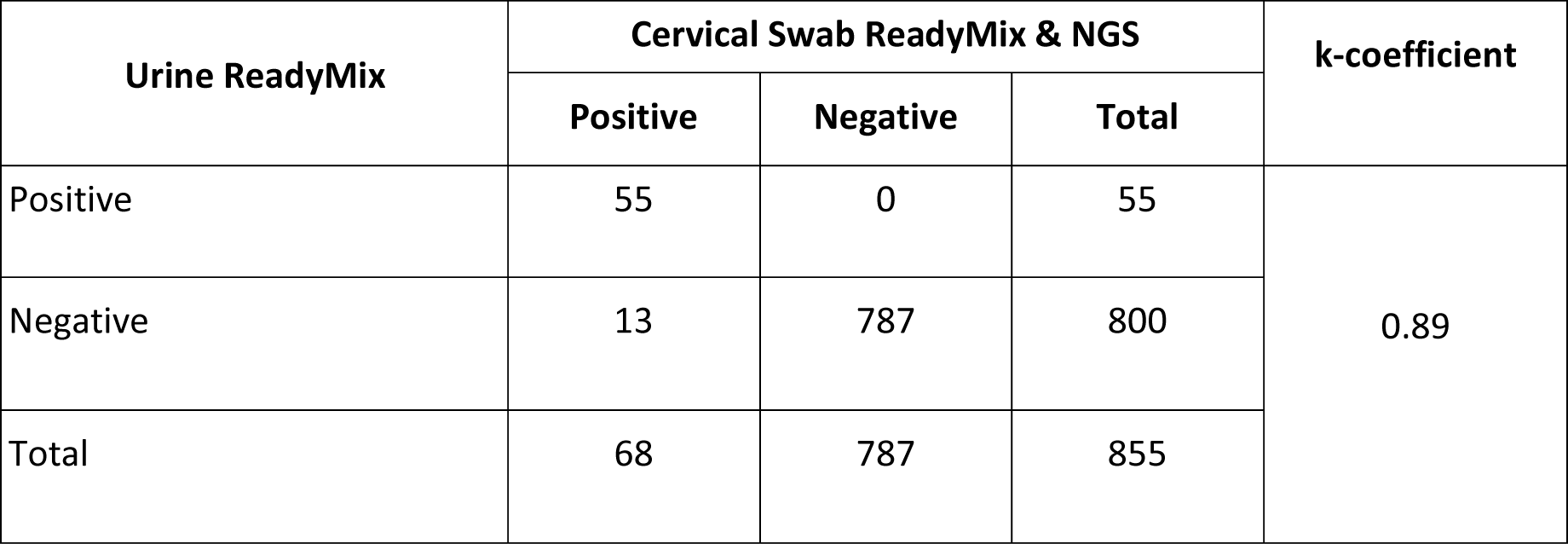
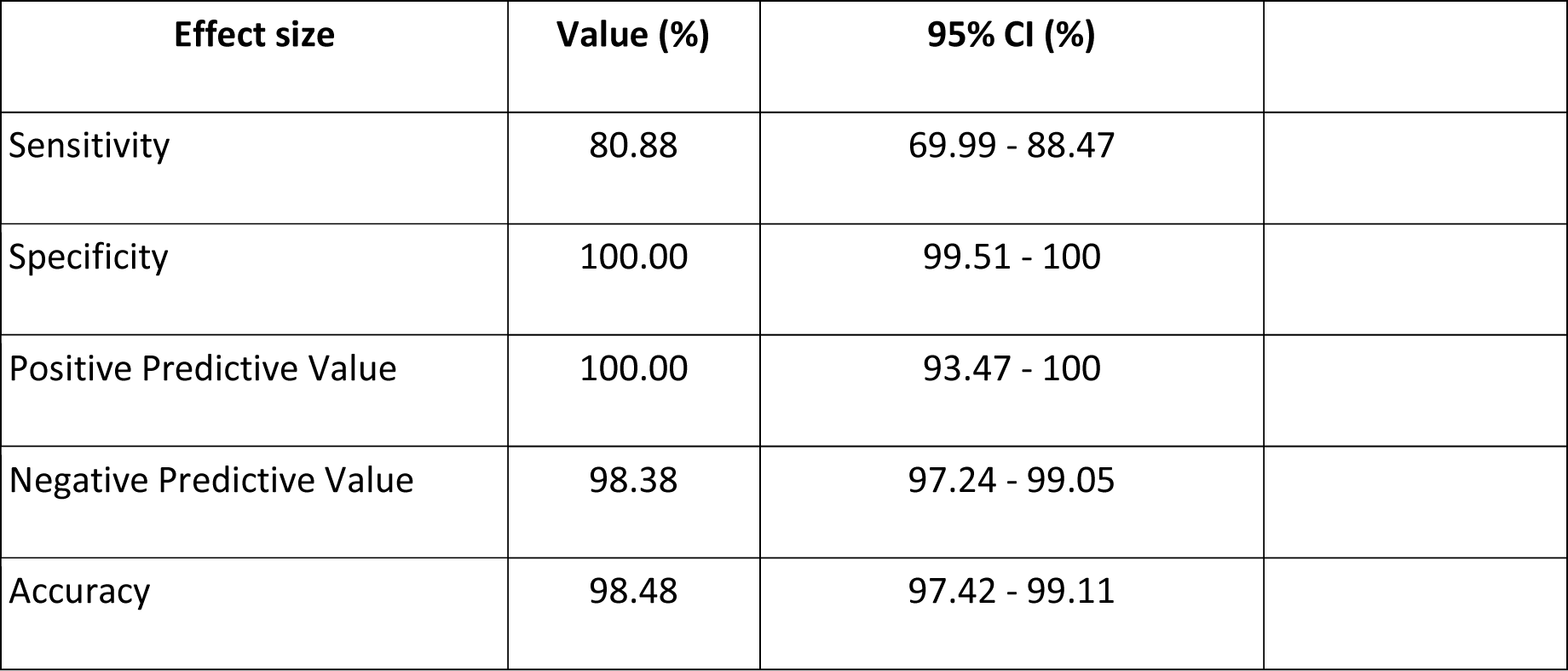
HPV detection using ReadyMix in urine samples versus cervical swabs and NGS.

| Urine ReadyMix | Cervical Swab ReadyMix & NGS |  |  | k-coefficient |
| --- | --- | --- | --- | --- |
|  | Positive | Negative | Total |  |
| Positive | 55 | 0 | 55 | 0.89 |
| Negative | 13 | 787 | 800 |  |
| Total | 68 | 787 | 855 |  |

| Effect size | Value (%) | 95% CI (%) |
| --- | --- | --- |
| Sensitivity | 80.88 | 69.99 - 88.47 |
| Specificity | 100.00 | 99.51 - 100 |
| Positive Predictive Value | 100.00 | 93.47 - 100 |
| Negative Predictive Value | 98.38 | 97.24 - 99.05 |
| Accuracy | 98.48 | 97.42 - 99.11 |

### Urine samples show a high sensitivity across low, moderate, and high cervical swab Ct value group

Upon analyzing the HPV’s Ct values of paired cervical swab and urine samples, a significant difference was observed between the two samples based on Wilcoxon matched-pairs signed rank test (p-value <0.0001) (Fig. 2A). The cervical swabs exhibited a lower median HPV’s Ct value (31.89 (8.41)) as compared to urine samples (35.69 (3.85)), resulting in 3.80 of median difference. The Ct value of IC detection also showed a similar pattern whereby cervical swabs produced a lower median (26.685 (3.82)) as compared to urine samples (30.69 (3.18)) (Fig. 2B).

**Figure 2.**
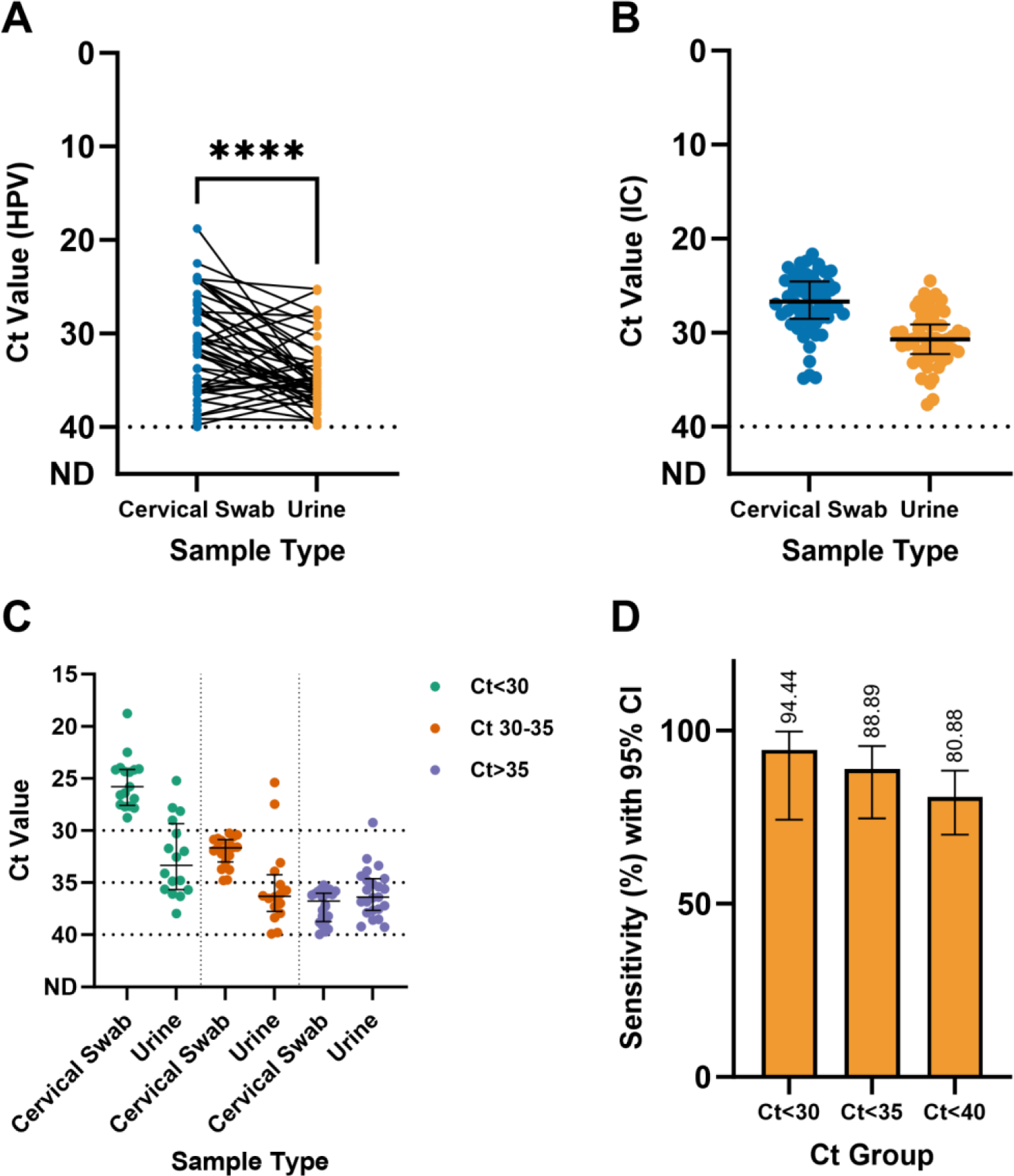
The Ct value of HPV detection in cervical swabs and urine samples was significantly different (Wilcoxon matched-pairs signed rank test, p-value: <0.0001), with cervical swabs tending to produce a lower Ct value (A). This finding was supported by a similar pattern of the Ct value of IC detection (B). However, the Ct value differences between cervical swabs and urine samples decrease as the sample’s Ct value increases (C), and the sensitivity of HPV detection in urine samples is maintained at a high level across all Ct values (D).

The HPV’s Ct value median differences of the two samples follow a pattern whereby the difference tends to be lower as the Ct values get higher. The low Ct group (Ct<30) of cervical swab samples had median Ct value of 26.08 (3.51) with the corresponding urine samples having a median Ct value of 34.11 (6.24) (Fig. 2C). The medium Ct group (Ct 30-35) had a median Ct value of 31.63 (1.53) in cervical swabs while the corresponding urine samples have a median Ct value of 36.26 (3.36) (Fig. 2C). The high Ct group (Ct>35), had cervical swab’s median Ct value of 36.76 (2.66) with the paired urine samples having a median of 36.39 (3.04) (Fig. 2C). Thus, the median difference between urine and cervical swabs for low, medium, and high Ct values were 8.03, 4.63, and 0.37, respectively. Despite the wider median gap at a lower Ct value and a tendency to produce higher Ct, the sensitivity of urine samples is maintained at a high level across all Ct groups, ranging from 80.88% positive percentage agreement when considering all samples to 99.44% positive percentage agreement when calculating for samples with Ct < 30 (Fig. 2D).

### hrHPV ReadyMix qPCR Kit and cobas^®^ 6800 HPV produced a comparable HPV prevalence pattern across different age groups and locations

The HPV prevalence of the study population was slightly higher when tested by ReadyMix as compared to cobas (Fig. 3A). ReadyMix showed an HPV prevalence of 6.62% and 6.28%, based on cervical swabs and urine respectively, while cobas showed an HPV prevalence of 5.94% and 5.82%, based on cervical swabs and urine respectively. As previously described, most of the additional positive samples were proven to be true-positives by NGS, suggesting the superior performance of the ReadyMix. However, we found a similar HPV prevalence shown by the two tests across different age groups and different locations.

**Figure 3.**
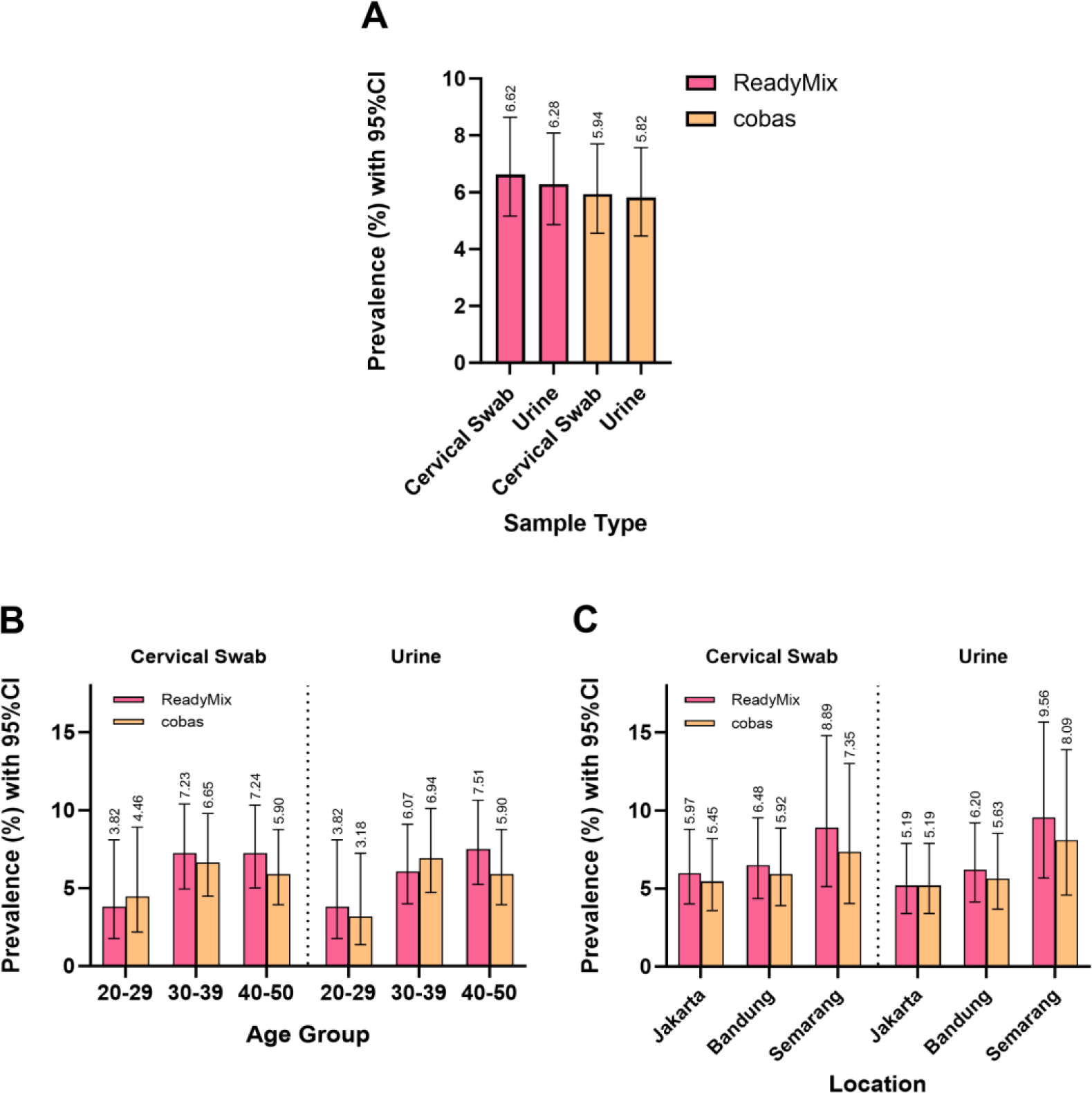
ReadyMix identified a higher HPV prevalence, as supported by NGS results, indicating increased sensitivity (A); This result slightly affected the prevalence distribution across different age groups, where ReadyMix found a higher prevalence in the age group 40-50 compared to 30-39, while cobas found the opposite (B). However, the two tests showed a similar trend, especially in discovering the age group 20- 29 to have the lowest prevalence among all groups. On the other hand, the prevalence distributions based on locations observed by ReadyMix and cobas were less affected, resulting in a similar pattern in both cervical swabs and urine samples (C).

The 20-29 age group had the lowest HPV prevalence according to both tests based on cervical swabs (ReadyMix: 3.82%; cobas: 4.46%) and urine (ReadyMix: 3.82%; cobas: 3.18%) (Fig. 3B). ReadyMix indicated a marginally higher prevalence in the age group of 40-50 (Cervical swab: 7.24%, Urine: 7.51%) compared to the 30-39 group (Cervical swab: 7.23%, Urine: 6.07%). Conversely, cobas showed a higher prevalence in the 30-39 group (Cervical swab: 6.65%, Urine: 6.94%) compared to the 40-50 group (Cervical swab: 5.90%, Urine: 5.90%) (Fig. 3B). This contrasting finding mostly result from several (5 cervical swabs and 5 urine) samples from the 40-50 group that were tested negative by cobas but were found to be HPV-positive by ReadyMix and confirmed by NGS (Supplementary Materials: Supplementary Table S1).

Comparing the prevalence across different locations, both tests show that Semarang had the highest HPV prevalence based on cervical swab samples (ReadyMix: 8.89%; cobas: 7.35%), followed by Bandung (ReadyMix: 6.48%; cobas: 5.92%), while Jakarta presented with the lowest HPV prevalence (ReadyMix: 5.97%; cobas: 5.45%) (Fig. 3C). This similar pattern was also observed in urine samples with Semarang’s HPV prevalence of 9.56% and 8.09% by ReadyMix and cobas respectively, followed by Bandung (ReadyMix: 6.20%; cobas: 5.63%) while Jakarta continued to have the lowest prevalence among the three locations (ReadyMix: 5.19%; cobas: 5.19%).

### Similar HPV proportion is observed in both urine samples and cervical swabs

Besides the similar HPV prevalence, the pattern of HPV proportion in urine samples was also comparable to that of cervical swabs, with HPV Others having the highest proportion (Cervical swab: 50.00%; Urine: 45.45%), followed by HPV52 (Cervical swab: 17.24%; Urine: 21.82%), HPV16 (Cervical swab: 15.52%; Urine: 10.91%), and HPV18 (Cervical swab: 5.17%; Urine: 7.27%) (Fig. 4A). There were also some coinfection cases, which occupied 12.07% and 14.55% of HPV infection cases detected in cervical swabs and urine samples respectively. Most of them were HPV52-HPV Others combination (Cervical swab: 57%; Urine: 50%) (Fig. 4B). The proportions of HPV16-HPV Others, HPV18-HPV Others, and HPV16-HPV52-HPV Others in cervical swabs are all equal at 14% while HPV16-HPV Others’ proportion (38%) is higher than HPV18-HPV Others (13%) in urine samples. The HPV16-HPV52-HPV Others case detected as HPV16-HPV Others contributed to this difference. A subset of coinfection samples was also confirmed by NGS (Supplementary Materials: Supplementary Table S1; Supplementary Fig.S3) with HPV31, HPV33, HPV51, HPV56, and HPV58 found, confirming the HPV Others detection by qPCR.

**Figure 4.**
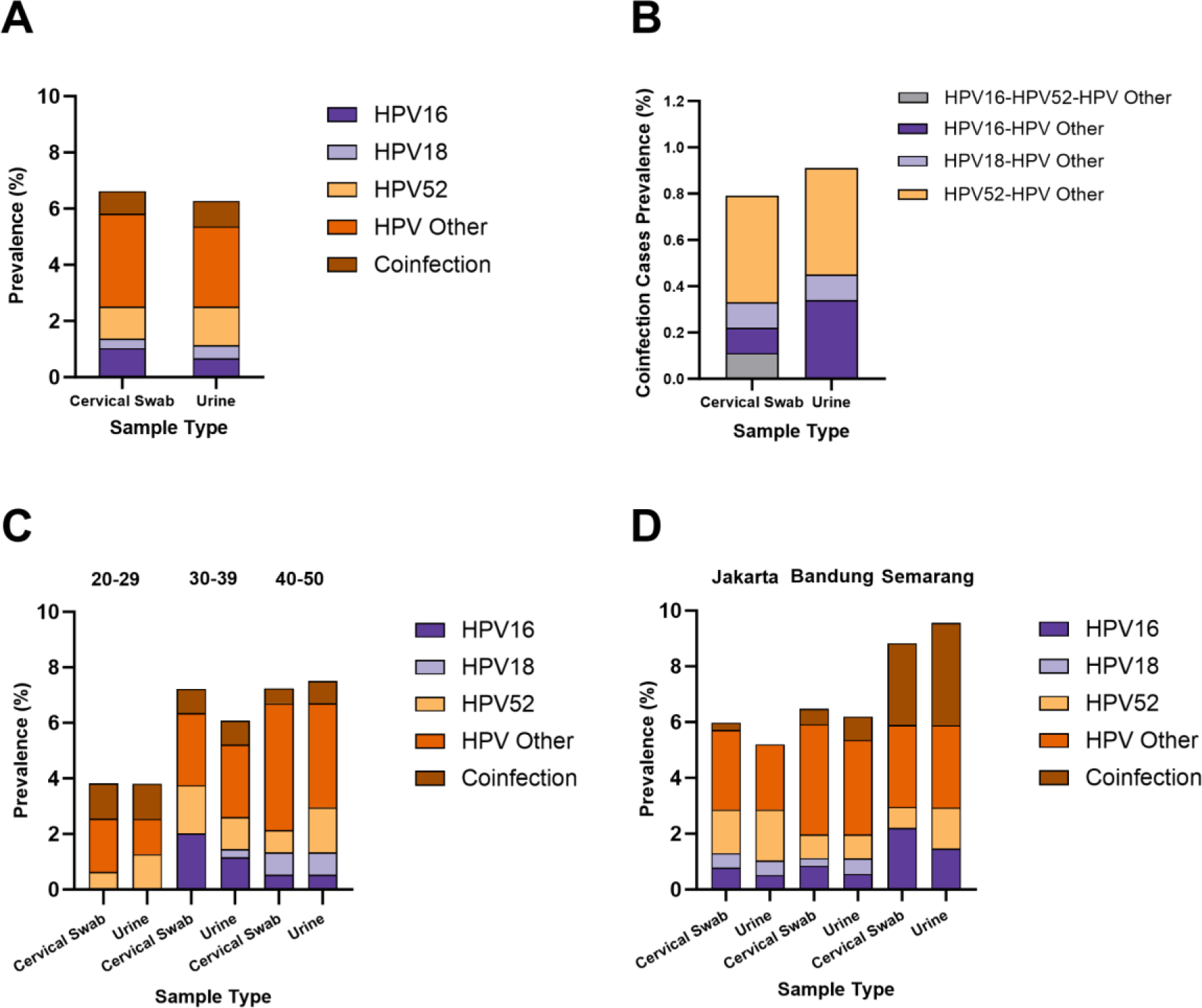
The HPV proportions of each HPV type/ group found in urine samples were comparable to cervical swabs (A). Urine samples also enable coinfection case detection, resulting in a pattern similar to cervical swabs (B). Generally, the HPV type/group proportions found in urine are similar to cervical swabs, across different age groups. A minor difference, where an HPV18 case found in the urine sample of the 30-39 group was confirmed by NGS (C). Except for the absence of coinfection cases in urine samples from Jakarta, the similarity in the pattern of HPV proportions between urine and cervical swabs was also observed in a location-based analysis (D).

The pattern of HPV proportion across different age groups by urine and cervical swabs share a considerable similarity (Fig. 4C). According to cervical swab samples, HPV Others dominated the HPV type detected, (age group 20-29: 50.00%, age group 30-39: 36%, and age group 40-50: 62.96%). The urine sample also showed an almost identical pattern (age group 30-39: 42.86%; age group 40-50: 50%), except for age group 20-29 where HPV Others share the same proportion as HPV52 and coinfection cases at 33%. In the age group of 30-39, HPV52 and HPV16 shared a similar proportion with 24.00% and 28.00% in cervical swabs, respectively, and both have 19.05% proportions in urine samples. A small proportion of HPV18 was detected in this group’s urine samples (4.76%), while none was found in cervical swabs. This presence of HPV genetic materials has been confirmed with NGS, showing that the detection of HPV18 was not a false-positive result. In the age group of 40-50, cervical swabs and urine displayed similar proportions of HPV16 (Cervical swab: 7.41%; Urine: 7.14%) and HPV18 (Cervical swab: 11.11%; Urine: 10.71%), while HPV52’s proportion was higher in urine samples (21.43%) compared to cervical swabs (11.11%). This minor variation could be attributed to the samples with discordant results between cervical swabs and urine.

Analyzing the HPV prevalence by location, HPV Others remained dominant, except in Semarang, where it shared the same proportion with coinfection cases (33.33%) as determined by cervical swab (Fig. 4D). Semarang, in fact, had the highest coinfection cases proportion in the urine sample (38.46%) when compared across all locations (Jakarta: 0.00%; Bandung: 13.64%). HPV52 holds the second largest proportion in Jakarta, in both cervical swabs (26.09%) and urine (35.00%), meanwhile, in Semarang, its proportion is the same as HPV16 at 15.38% in urine or lower when using cervical swabs (HPV52: 8.33%; HPV16: 25.00%). Both cervical swabs and urine displayed some small HPV18 proportions in Jakarta (Cervical swabs: 8.7%; Urine: 10%) and Bandung (Cervical swab: 4.35%; Urine: 9.09%), while there were none detected in Semarang. A single coinfection case was found in Jakarta based on cervical swabs (4.35%), while none was detected in urine samples. In Bandung, cervical swabs and urine showed a similar HPV52 proportion (Cervical swabs: 13.04%; Urine: 13.64%) while the HPV16 proportion is higher in the cervical swabs (13.04%) than in urine (9.09%).

### HPV16 and HPV18 genotyping results by hrHPV ReadyMix qPCR Kit are proportional to cobas^®^ 6800 HPV

To further validate the performance of ReadyMix, we compared the genotyping of HPV16 and HPV18 capability to cobas. The HPV16 proportion out of the positive HPV samples was found to be higher in cervical swabs (ReadyMix: 16.67%; cobas: 19.30%) as compared to urine samples (ReadyMix: 14.29%; cobas: 13.21%) (Fig. 5A, Supplementary Materials: Supplementary Table S4). The distribution of HPV16 across different age groups is consistent among all samples tested by ReadyMix and cobas with the highest prevalence observed in the age group of 30-39 (Cervical swab ReadyMix: 27.59%, cobas: 32.00%; Urine ReadyMix: 25.00%, cobas: 24.00%), followed by the 40-50 age group (Cervical swab ReadyMix: 10.34%, cobas: 12.00%; Urine ReadyMix: 9.68%, cobas: 4.35%) while no HPV16 found in the age group of 20-29 (Fig. 5B, Supplementary Materials: Supplementary Table S4).

**Figure 5.**
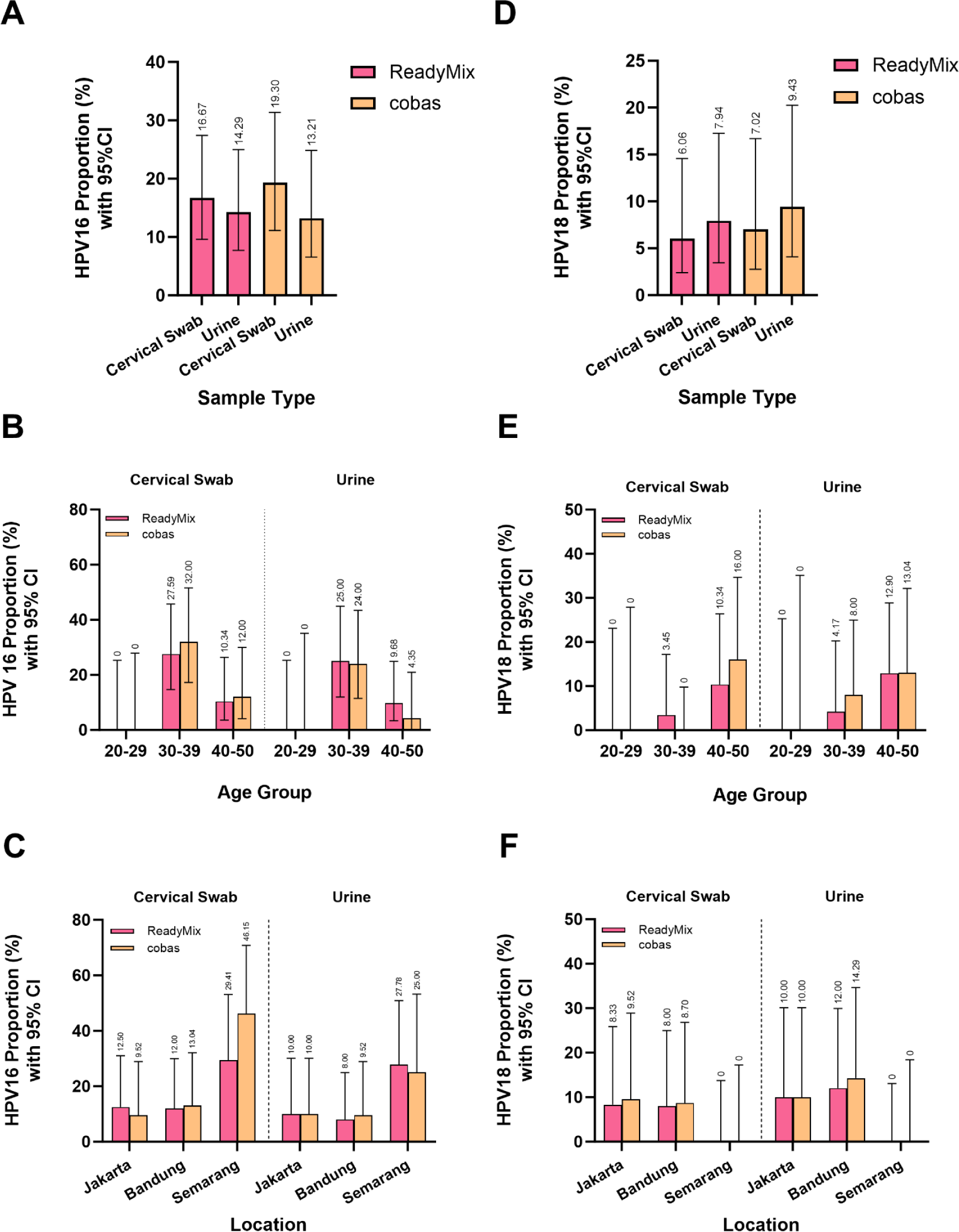
The pattern of HPV16 proportions among positive cases resulting from ReadyMix was similar to cobas, where cervical swabs had a higher HPV16 proportion compared to urine (A). The similarity was also observed when grouping the subjects into different age (B) and location categories (C). On the other hand, the overall proportion of HPV18 found in urine samples was slightly higher than in cervical swabs, as shown by both ReadyMix and cobas (D). The pattern was also observed when analyzing based on age groups, except for cobas result in age group 40-50 where cervical swab has a slightly higher HPV18 proportion (E). The HPV18 proportion across different locations, as detected by ReadyMix and cobas, remained consistent, even in the presence of a minor pattern discrepancy between cervical swabs and urine samples (F).

When looking at HPV16 prevalence from the perspective of location origins, cobas detected a higher proportion of HPV16 (46.15%) in cervical swabs samples when compared to results detected by ReadyMix (29.41%) in Semarang (Fig. 5C). The total number of HPV-positive samples detected with cobas were smaller when compared to ReadyMix (ReadyMix: 17 HPV-positives; cobas: 13 HPV-positives) while the HPV16-positive samples detected with both ReadyMix and cobas were similar (ReadyMix: 5 HPV16-positives; cobas: 6 HPV16-positives), resulting in the difference in HPV16 prevalence. On the other hand, the urine samples exhibited a more consistent results with 27.78% and 25.00% of HPV16 proportion when tested with ReadyMix and cobas respectively. Cervical swab samples tested with ReadyMix gave similar proportions of HPV16 in Jakarta (12.50%) and Bandung (12.00%) while a slightly lower HPV16 proportion in Jakarta was obtained when tested using cobas (Jakarta: 9.52%; Bandung: 13.04%). On the other hand, Bandung had a slightly lower HPV16 proportion as compared to Jakarta when urine samples were tested with ReadyMix (Jakarta: 10.00%; Bandung: 8.00%) and cobas (Jakarta: 10.00%; Bandung: 9.52%).

As for HPV18 prevalence, both tests showed a higher prevalence in urine samples (ReadyMix: 7.94%; cobas: 9.43%) when compared to cervical swab samples (ReadyMix: 6.06%; cobas: 7.02%) (Fig. 5D). The HPV18 proportion out of the positive HPV samples, in general, was increased with increasing age group, except for the age group 30-39 where no samples tested HPV18 positive (Fig. 5E). The age group of 40-50 had the highest HPV18 positive proportion of 10.34% (ReadyMix) and 16.00% (cobas) on the cervical swab samples and 12.90% (ReadyMix) and 13.04% (cobas) on the urine samples. There were no samples with HPV18 tested positive in the 20-29 group.

Location-based HPV positive proportion analysis indicated an agreement between the two tests on the slightly higher HPV18 proportion in Jakarta (ReadyMix: 8.33%; cobas: 9.52%) when compared to Bandung (ReadyMix: 8.00%; cobas: 8.70%) based on cervical swabs (Fig. 5F). On the other hand, urine samples showed a slightly higher HPV18 proportion found in Bandung (ReadyMix: 12.00%; cobas: 14.29%) compared to Jakarta (ReadyMix: 10%; cobas: 10%). There were no samples in Semarang that tested HPV18 positive.

## DISCUSSION

Our study demonstrates the good performance of the ReadyMix Kit in detecting HPV using both standard cervical swab samples and urine samples, achieving a diagnostic accuracy of 99.65% and 98.48% respectively. These results were also validated by NGS in which the dispute samples were sequenced to confirm the presence of HPV genetic materials. We also validated that the prevalence data of the high-risk HPV using urine samples are similar to those of the cervical swab samples, indicating ReadyMix Kit’s reliability in cervical cancer screening.

The convenience of using urine as a sample for HPV detection has garnered interest as an alternative to the conventional cervical swabbing method. The cervical examination is frequently seen as an uncomfortable and embarrassing procedure, particularly among women in emerging countries, restraining them from getting screened.^20,21^ Nishimura et al. (2021) have indicated that self-sampling is more acceptable as it protects privacy and reduces anxiety.^22^ Furthermore, Zhao et al. (2022) noted that women showed higher confidence in correctly collecting urine samples rather than cervical samples.^7^ Besides, the usage of self-collected urine sample diminishes the need of skilled personnel and special supporting equipment required for cervical swab sample collection.

Urine samples have been reported to show a promising performance compared to conventional cervical swab samples for the detection of HPV.^9–11^ Unlike its preference for the cervix and other anogenital areas, HPV demonstrates no inherent affinity for the urinary tract. Consequently, the detection of HPV in urine likely signifies secondary exfoliation originating from cervical or other anogenital lesions.^23^ Therefore, utilizing first-void urine is crucial to acquire a greater quantity of exfoliated cells, which are likely to have a higher concentration of HPV DNA.^9^ In this study, we opted to collect first-void urine at any time of the day for participants’ convenience as there was no disparity in HPV DNA concentrations between morning and subsequent urine samples as long as they were obtained within the first-void phase.^24^ A comprehensive analysis of multiple studies revealed that the combined sensitivity of high-risk HPV detection in urine samples was 77%, with a specificity of 88%, when compared to cervical samples.^9^ We observed a higher sensitivity and specificity at 80.88% and 100%, respectively when tested using ReadyMix. The increased value was most likely influenced by NGS confirmation, which verifies initially false-positive results to be true positives. Furthermore, ReadyMix showed lower rates of invalid results in urine samples compared to cobas. In addition to the satisfactory detection rates, we illustrated that the HPV genotypes detected in urine are comparable to those found in cervical swabs. This finding aligns with the observations of Sahasrabuddhe et al., who noted a notably high concordance rate (97.1- 100%) between HPV genotypes identified in urine samples and vulvar sampling, and that sampled from cervical lesion tissue.^23^ This result highlights the potential of HPV detection in urine by ReadyMix as a reliable screening method.

ReadyMix’s good performance can also be shown in its similar HPV16 and HPV18 genotyping results with cobas. This confirms the proficiency of ReadyMix in accurately determining the HPV16 and HPV18-positive cases, providing clinicians with valuable information to guide subsequent measures. The 2019 ASCCP Risk-Based Management Consensus Guidelines suggests an additional evaluation such as colposcopy to be performed in HPV16 and HPV18-positive patients, even when cytology results are negative.^25^ HPV52 genotyping as an additional genotyping capability of ReadyMix also helps in covering the HPV type with the highest proportion among high-risk HPV detected in Indonesia.^26^ Our study found a higher proportion of HPV52 compared to HPV16 and HPV18 in both cervical swabs (HPV52: 17.24%; HPV16: 15.52%; HPV18: 5.17%) and urine (HPV52: 21.82%; HPV16: 10.91%; HPV18: 7.27%). The result was in line with a previous study where HPV52, HPV16, and HPV18 comprised 23.2%, 18.0%, and 16.1% of the high-risk HPV-positive cases in Indonesia.^26^ Despite the 15-year gap, our study’s findings continue to emphasize the necessity of a vaccine covering the HPV52 type, in addition to HPV16 and HPV18, for the Indonesian population.

The screening data by ReadyMix shows an HPV prevalence of 6.28-6.62%, slightly higher than a previous study conducted in Indonesia with a total HPV prevalence of 5.2%, with high-risk HPV contributing to 4.4% prevalence.^13^ However, the prevalence is still lower than the worldwide prevalence of high-risk HPV infection which is 10.4%.^27^ Unlike most studies where the HPV prevalence decreases with increasing age,^28^ we found a lower HPV prevalence in the lowest age group of 20-29 (Cervical swab: 3.82%; Urine: 3.82%) as compared to age group 30-39 (Cervical swab: 7.23%; Urine: 6.07%) and age group 40-50 (Cervical swab: 7.24%; Urine: 7.51%). Vet et al. (2008) also observed that there was no decline in HPV prevalence in the subjects of the older age group from Jakarta and Tasikmalaya.^26^ Additionally, the increasing prevalence in the older age groups was also found by Yang et al. (2020) in China, resulting in the highest overall HPV prevalence held by age group ≥50 (10.53%), followed by 30-49 (8.29%), and the lowest held by age group <30 (7.30%).^29^ Several factors that may account for this phenomenon include a lower number of representatives in the young age group caused by the randomized subject screening methodology of this study as well as potentially reduced immunological functions of the older group, making them more susceptible to persistent HPV infection.^29^ Based on these data, we suggest an increased effort in the diagnosis of HPV infection in older women as they have a higher risk of cervical cancer progression and lower protection derived from vaccination^30^.

We found HPV prevalence to be the highest in Semarang (Cervical swab: 8.82%; Urine: 9.56%), followed by Bandung (Cervical swab: 6.48%; Urine: 6.20%), and lastly Jakarta (Cervical swab 5.97%; Urine: 5.19%). Here we can see that Jakarta, the biggest metropolitan city, had the lowest HPV prevalence as compared to Semarang, a metropolitan city with a larger rural area. Bandung, positioned with a metropolitan rating between the two cities,^31^ also exhibits a prevalence value that falls between the prevalence of the two aforementioned cities. Despite many lifestyle factors influencing HPV prevalence in a community,^29^ Sabeena et al. (2017) discovered that the infection rate in rural women is higher than in those who lived in urban areas due to lower access to cancer screening and oncology care.^32^ This is a continuing problem for emerging countries due to the inequality of access to healthcare services.

In our study, we identified multiple HPV positive signals in 7 out of 58 HPV-positive cervical swab samples (12.07%) and 8 out of 55 HPV-positive urine samples (14.54%) (HPV16/HPV18/HPV52/HPV Other). Additionally, our NGS data (Supplementary Materials: Supplementary Table S1) indicate that some samples positive for HPV Other may harbor multiple HPV types. It is well-documented that individuals positive for HPV often exhibit infections with multiple HPV types^33–35^. However, the precise implications of this phenomenon for the onset and progression of cervical cancer remain elusive. While some studies suggest a lack of clear correlation between multiple HPV infections and cervical histology,^36,37^ others, propose that multiple infections of specific types may synergistically elevate the risk of cervical carcinoma.^38,39^ However, some additional studies indicate that the disease risk is only similar to the sum of the estimated risk of individual types, with little evidence of synergistic interactions.^34,40^ Considering that certain HPV types are inherently more carcinogenic than others,^41–43^ we suggest that prioritizing the detection of these more carcinogenic HPV types holds greater utility in initial screening protocols than attempting to differentiate among all HPV types within an infection.

Our study to evaluate the usage of urine samples for cervical cancer screening using the hrHPV ReadyMix Kit has several strengths. In addition to having a high number of participants across multiple age groups, we also enrolled subjects from three major cities with varying metropolitan ratings and likely different lifestyles.^30^ This is particularly important to ensure that the HPV detection method through urine samples is applicable to the Indonesian population. Our study design also ensured that the paired urine samples and cervical swabs were collected on the same day to reduce the variability of the result that might arise from different sampling times.

Additionally, urine samples were verified to be collected before the cervical swab to reduce bias from additional cell debris that might come from the swabbing procedure. We also implemented NGS to confirm the potential false-positive qPCR results to check the presence of HPV in both cervical swabs and urine samples, which resulted in the confirmation of HPV infection that was not detected by cobas. However, the main weakness of our study is that we did not perform any cervical visual inspection to confirm the subject’s disease status and its relevance to qPCR results. These limitations should be overcome by future studies to obtain a better perspective of high-risk HPV condition.

## ACKNOWLEDGEMENTS

We want to express our heartfelt thanks to all the dedicated healthcare personnel and participants who made this research possible through their hard work and voluntary sample contributions. Our appreciation extends to Cipto Mangunkusumo National Hospital for providing essential facilities for sample analysis and to Nusantics, PT Riset Nusantara Genetika for providing the ReadyMix kits and their valuable sequencing resources. We are grateful to PT. Bio Farma for their financial support, which greatly assisted in our study. We would like to acknowledge dr. Sri Harsi Teteki, dr. Indriastuti Soetomo, and Dicky Mahardika for their vital role in managing the project. These contributions were crucial in our research, and we are thankful for the collaboration and support that made this study successful.

## DATA AVAILABILITY

- Data description: This dataset comprises raw data collected during a clinical trial assessing the effectiveness of urinary high-risk HPV DNA detection to improve cervical cancer screening in developing countries. The trial aimed to evaluate the feasibility and accuracy of a non-invasive urine collection method paired with a novel multiplex qPCR test developed by a local Indonesian company. The dataset is provided in CSV format. The manuscript stemming from this clinical trial will be submitted to the Journal of Clinical Microbiology, American Society for Microbiology.
- Name(s) of the repository/repositories: FigShare (https://figshare.com/)
- Digital object identifiers (DOIs) or accession numbers: https://doi.org/10.6084/m9.figshare.26199875.v1

## Declaration of interest

N.S.I., R.U., S.M.K., and M.R.R. are employees of Nusantics, PT Riset Nusantara Genetika. R.U. has financial holdings in Nusantics, PT Riset Nusantara Genetika. N.N. and R.M.S are employees at PT Bio Farma. The remaining authors declare that they have no conflict of interest.

## Submission declarations and verification

This article is not currently under consideration for publication elsewhere. The publication of this article has received approval from all authors and has been implicitly or explicitly endorsed by the responsible authorities at the institution where the work was conducted.

## Role of the Funding Source

This study was funded by PT Bio Farma in terms of subject recruitment, sample processing, and human resources. PT Bio Farma has a role in the study design, subject recruitment, and decision to submit the paper for publication. The development of the hrHPV ReadyMix kit and the NGS method were funded by Nusantics, PT Riset Nusantara Genetika, which also has a role in the study design, data collection, analysis, interpretation, report writing, and the decision to submit the paper for publication.

## Author’s contribution

R.U., D.W., A., and N.N. designed the study. R.U. designed the test kit. D.W., R.W., and R.M.S. recruited the subjects. N.S.I., D.W., R.W., and R.M.S managed subjects’ data. N.S.I., R.U., D.W., R.W., and I.S.W., analyzed and interpreted the data. N.S.I. and R.W. performed laboratory experiments. S.M.K and M.R.R performed NGS experiments, analyzed, and interpreted NGS data. N.S.I and R.U drafted the first version of the manuscript. N.N and A. provided valuable suggestions to the manuscript. All authors read the manuscript, provided feedback, and approved the submission.

## Supplementary Materials

### 1. Supplementary Figures

**Supplementary Fig S1.**
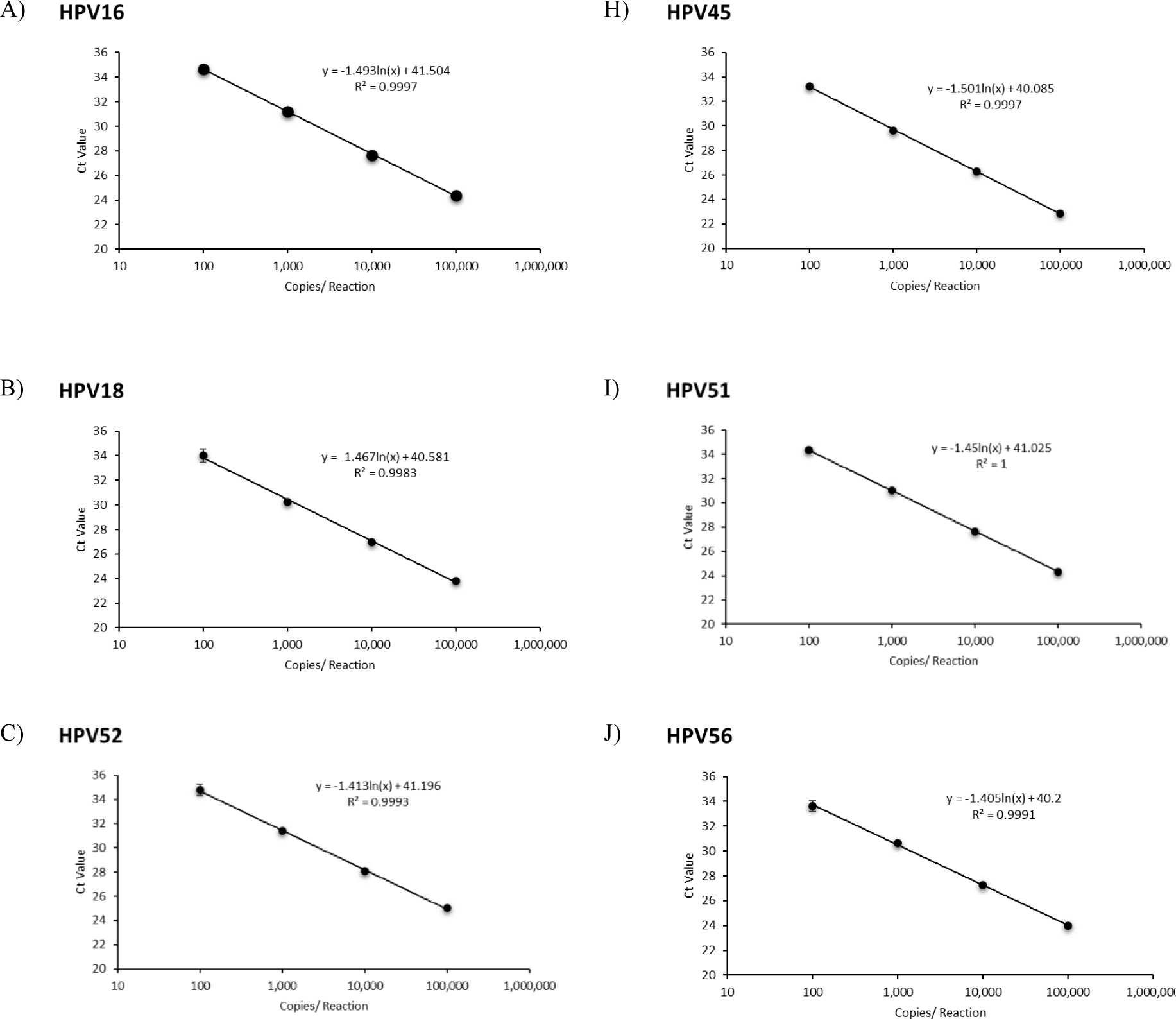
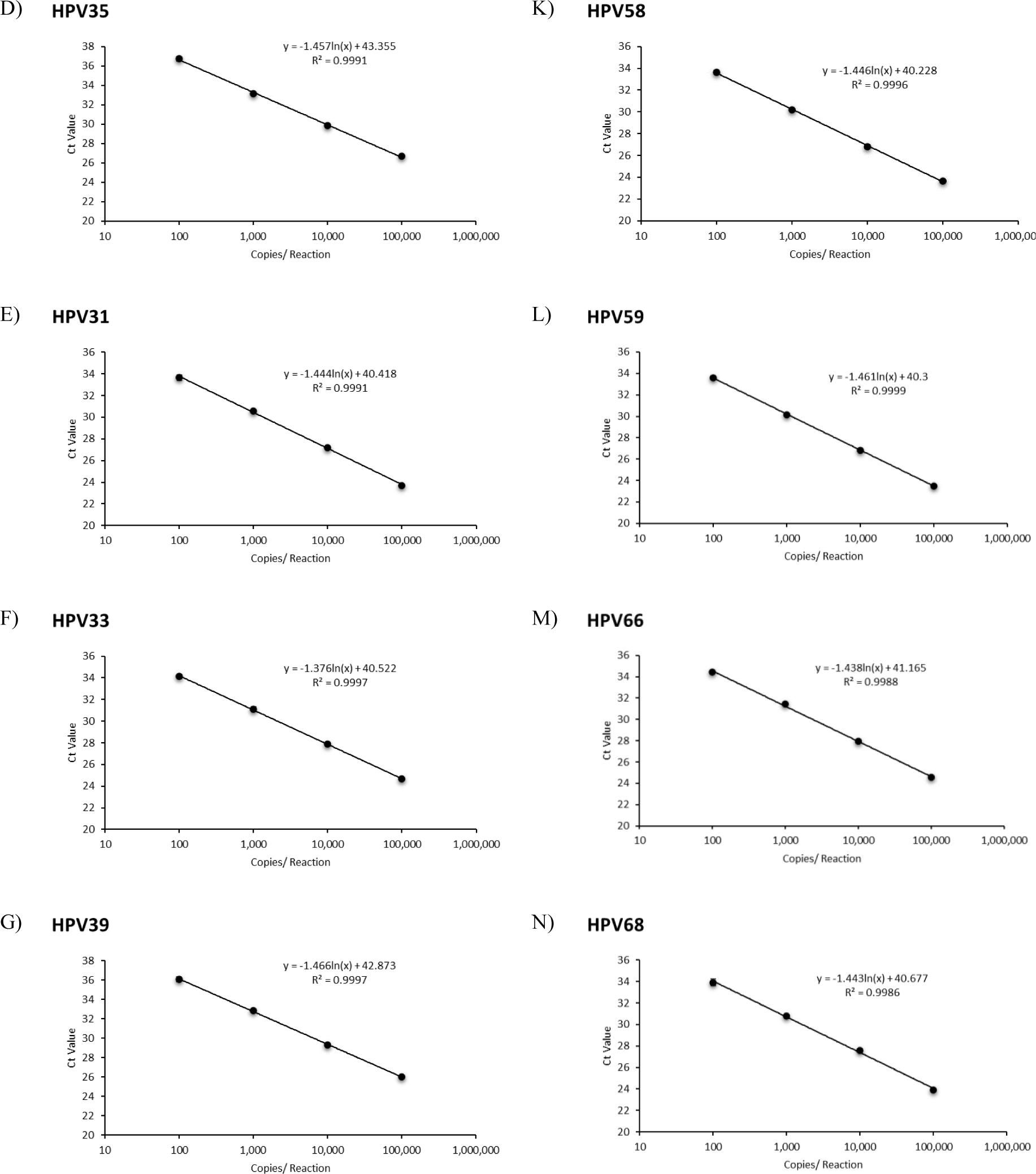
Linearity assay of ReadyMix in detecting 14 hrHPV types using HPV synthetic DNA as template ranging from 100 to 100,000 GE copies/reaction. A) HPV16; B) HPV18; C) HPV52; D) HPV35; E) HPV31; F) HPV33; G) HPV39; H) HPV45; I) HPV51; J) HPV56; K) HPV58; L) HPV59; M) HPV66; N) HPV68. This assay demonstrated excellent efficiency with a range between 90-110% and R^2^ ∼1.

**Supplementary Fig S2.**
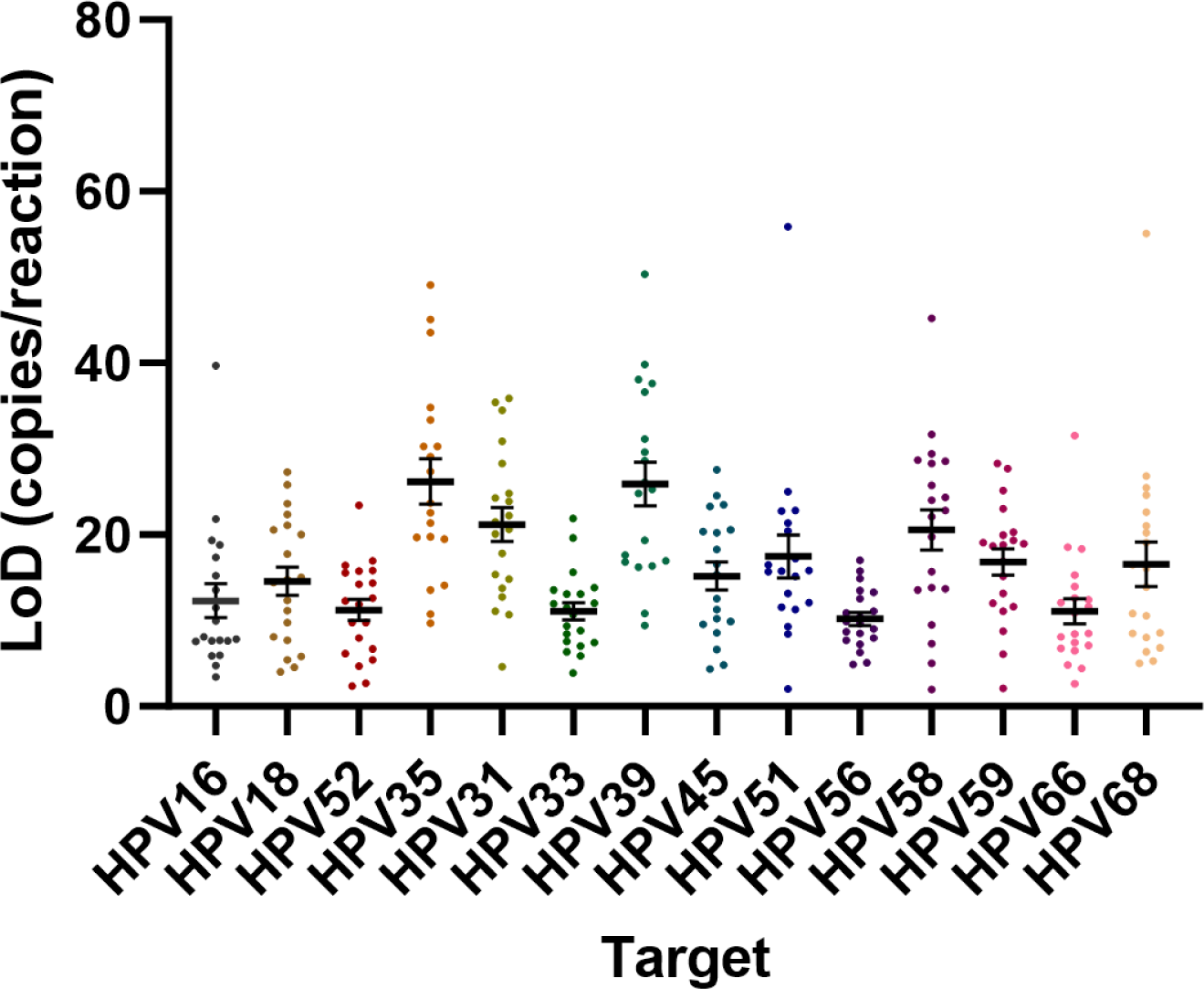
Limit of Detection (LoD) of ReadyMix for each hrHPV target ranging from 10-28 copies/20 uL qPCR reaction

**Supplementary Fig S3.**
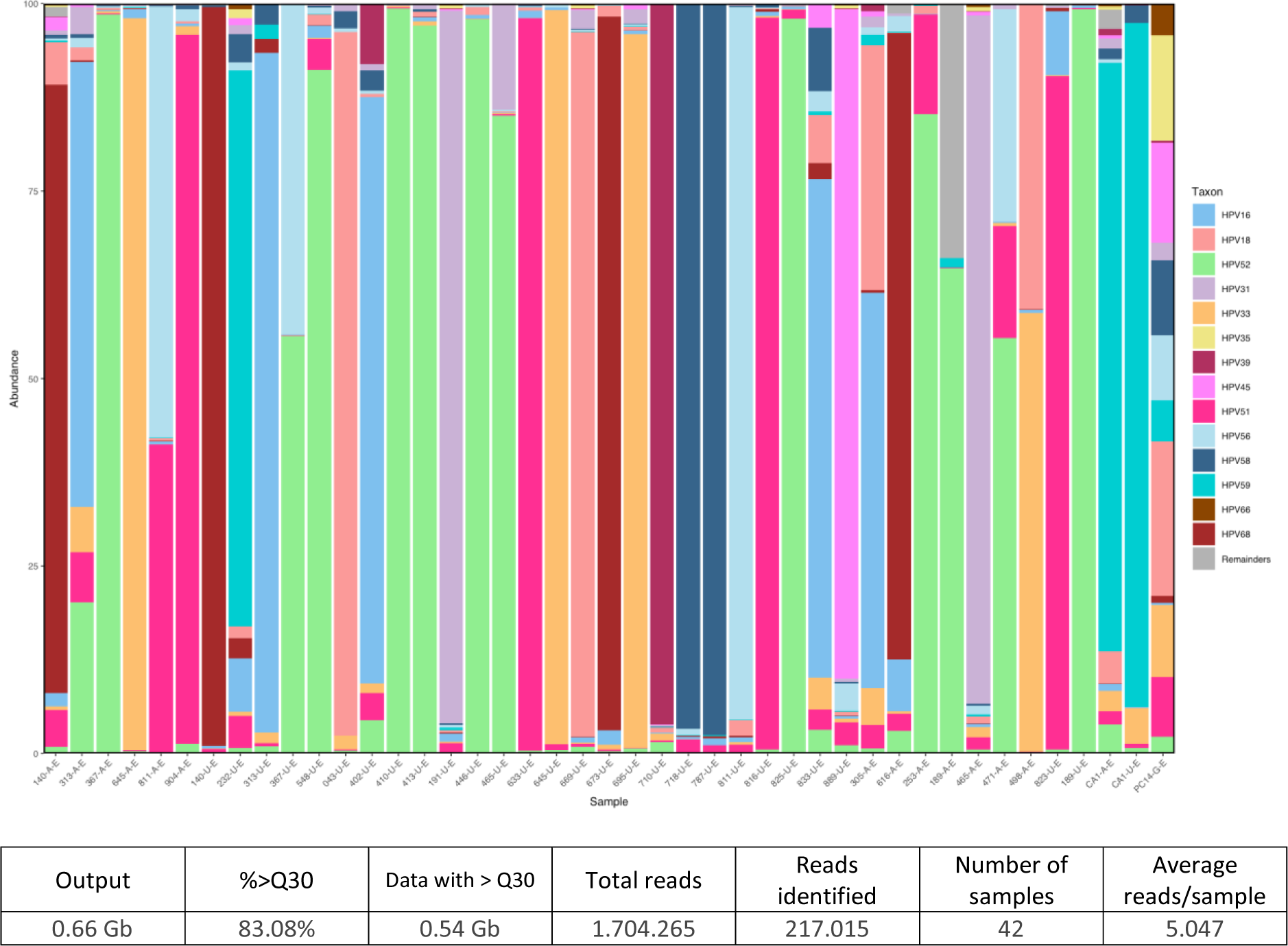
Stacked taxa bar plot showing the relative abundance of HPV type found in the sample

### 2. Supplementary Tables

**Supplementary Table S1.**
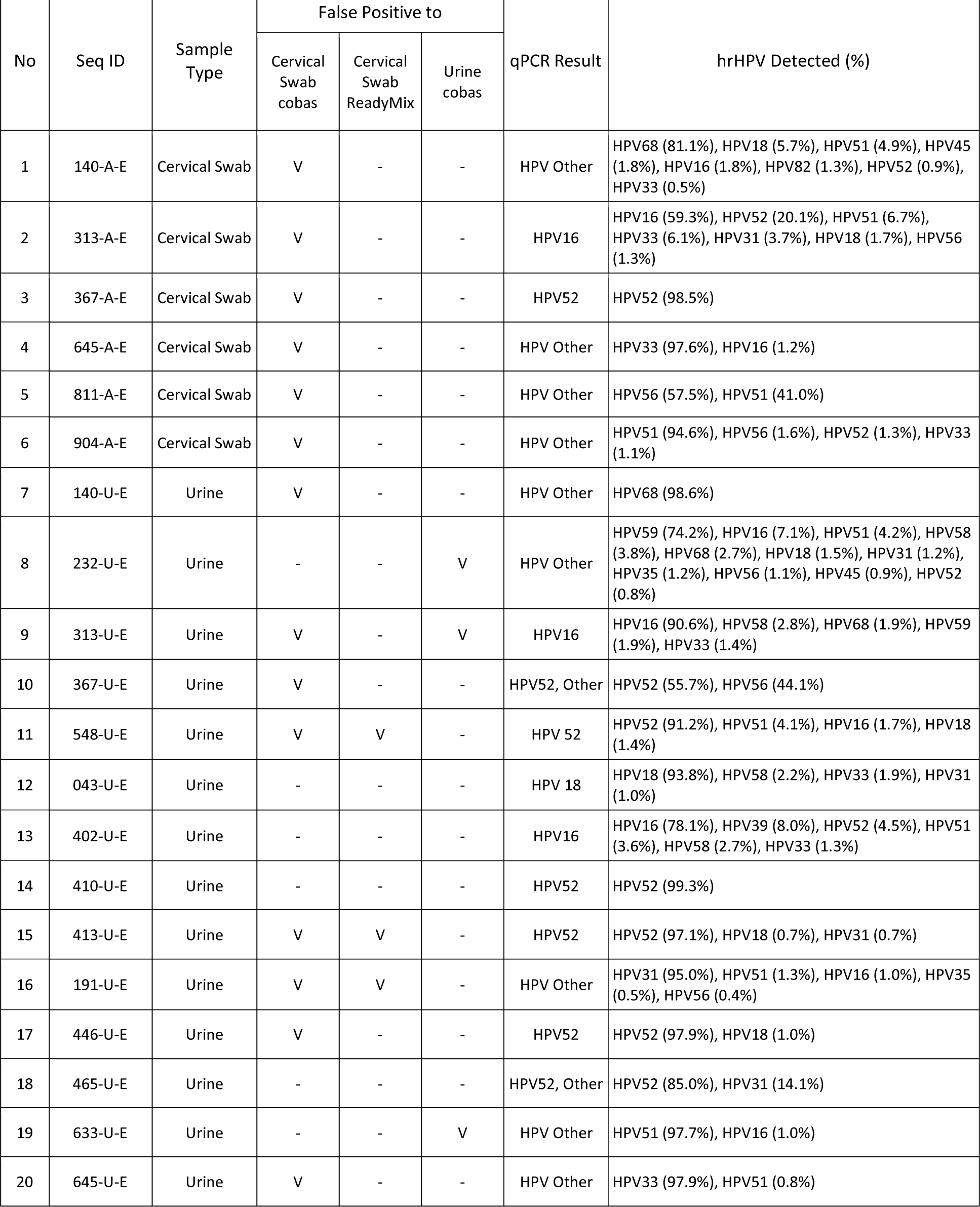
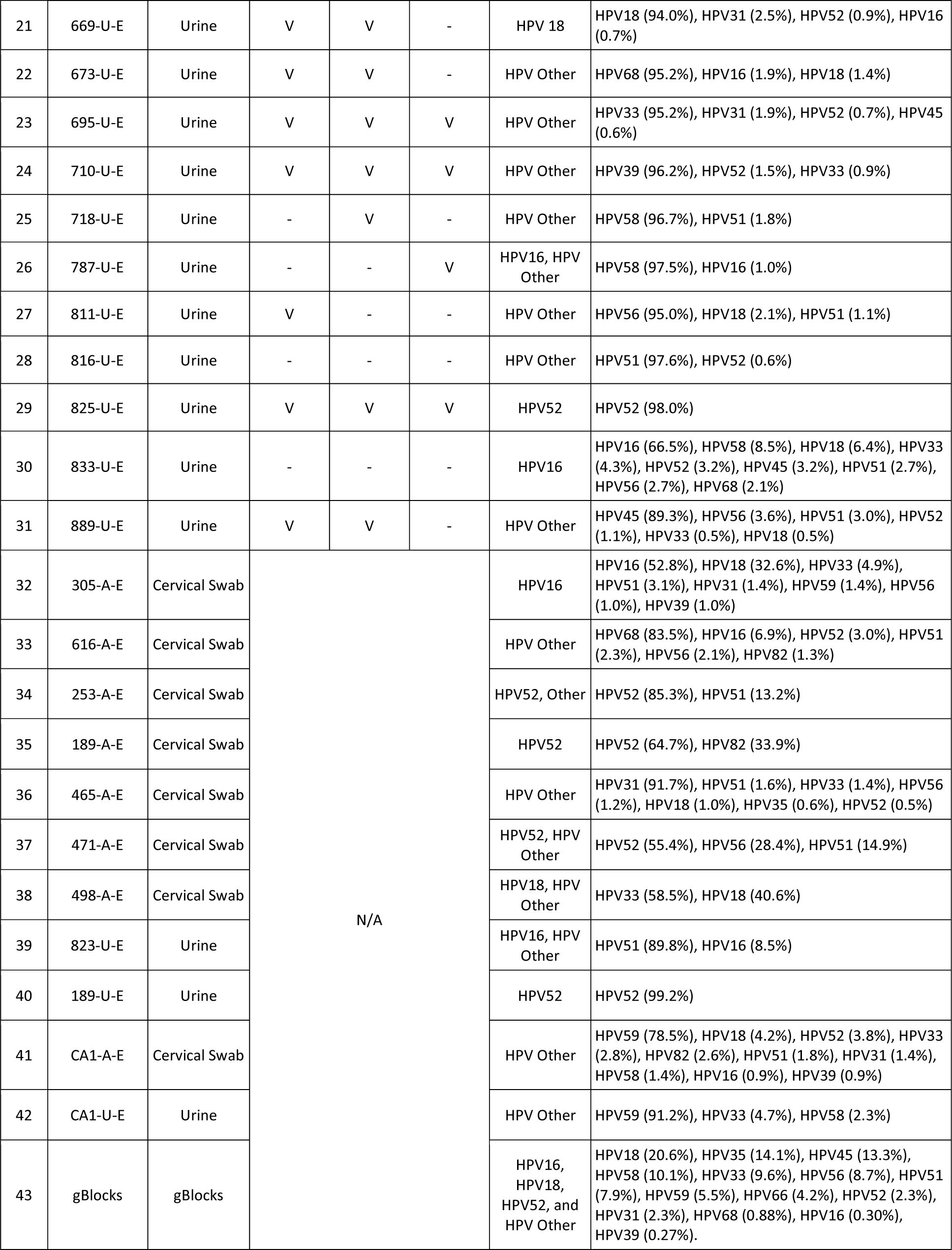
The results of NGS run to investigate the presence of hrHPV in the samples based on E6-E7 amplicon sequencing.

**Supplementary Table S2.**
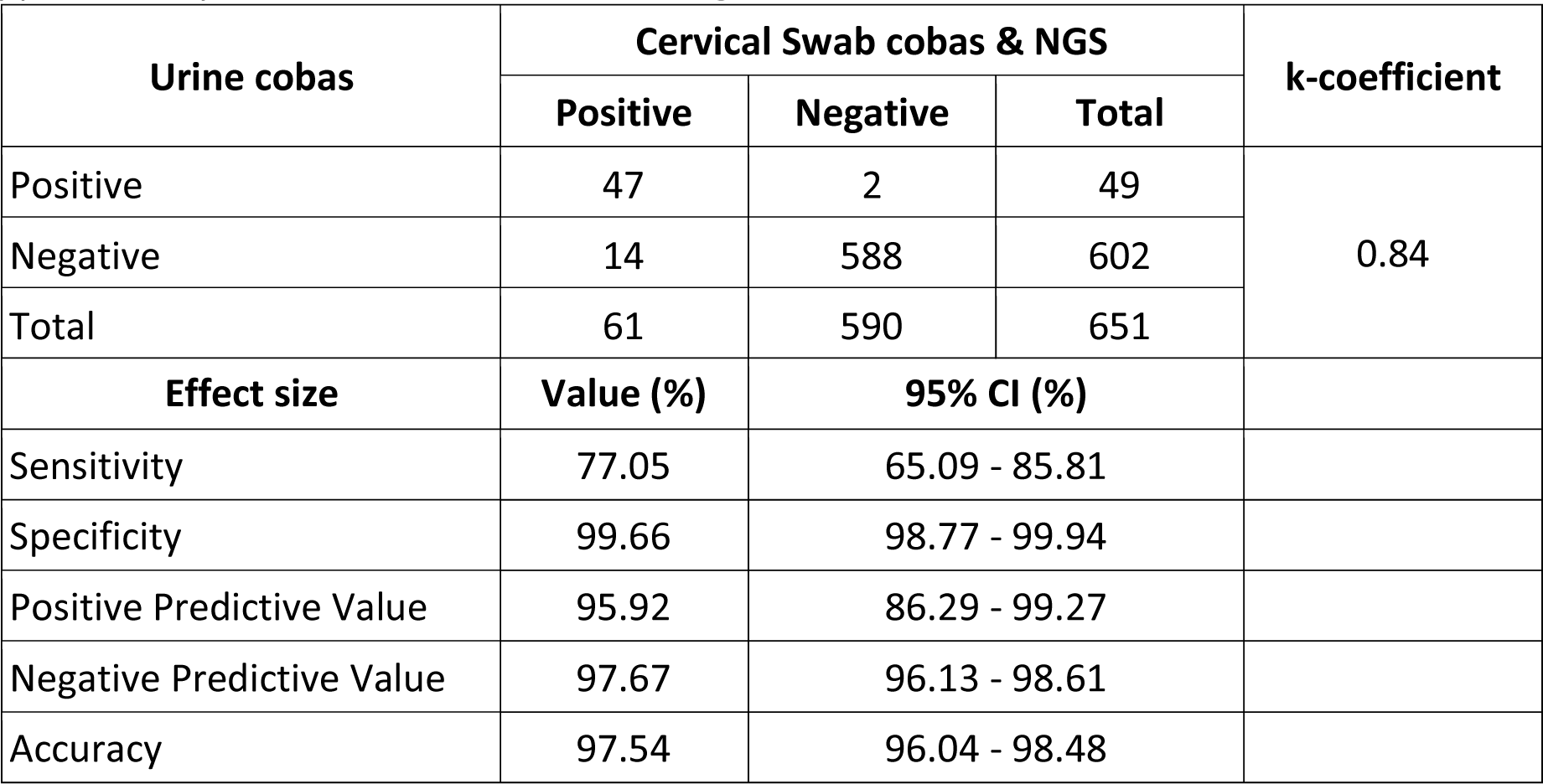
HPV detection using cobas in urine versus cervical swab and NGS.

**Supplementary Table S3.**
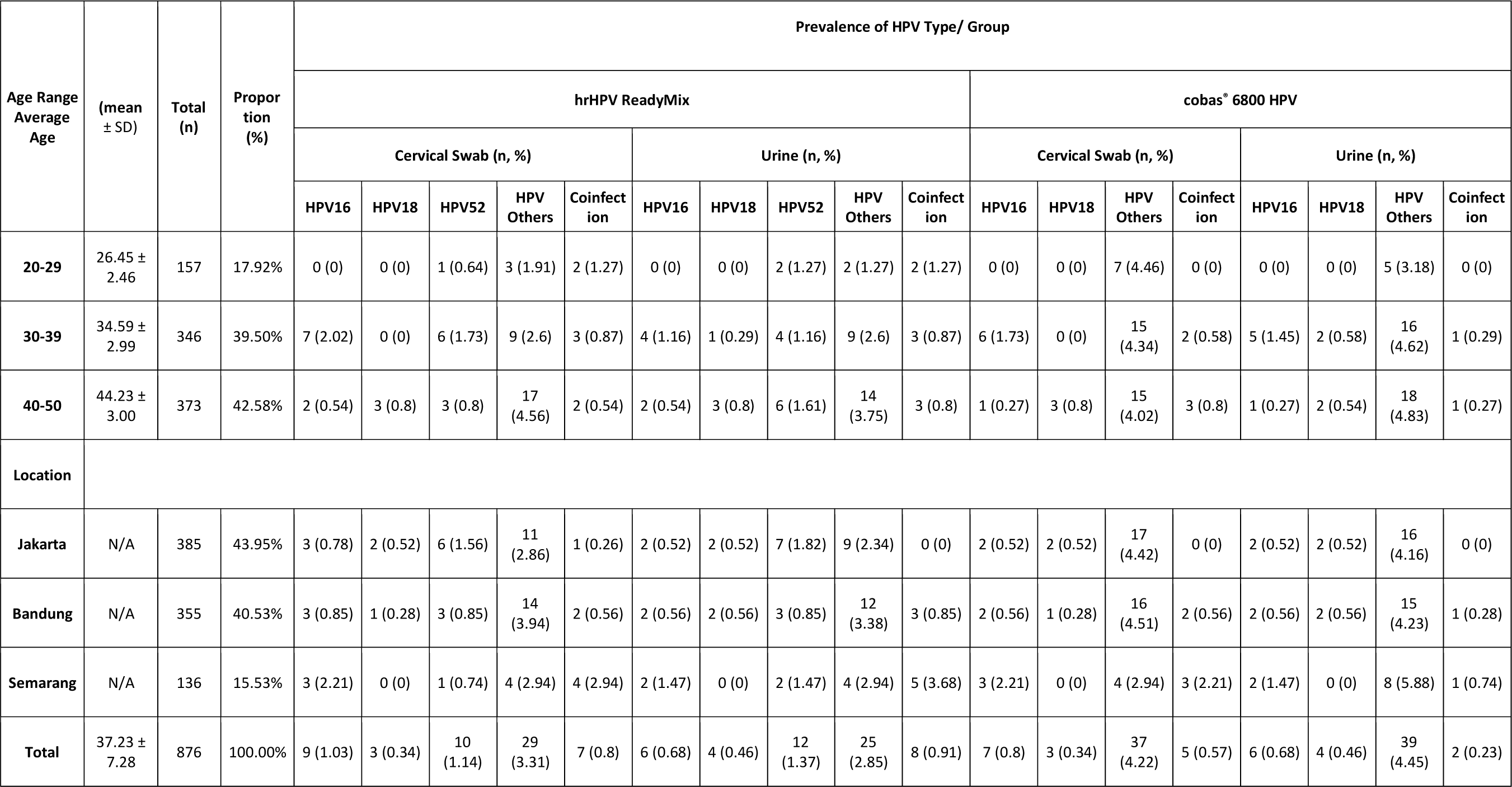
Demographics of the study population.

**Supplementary Table S4.**
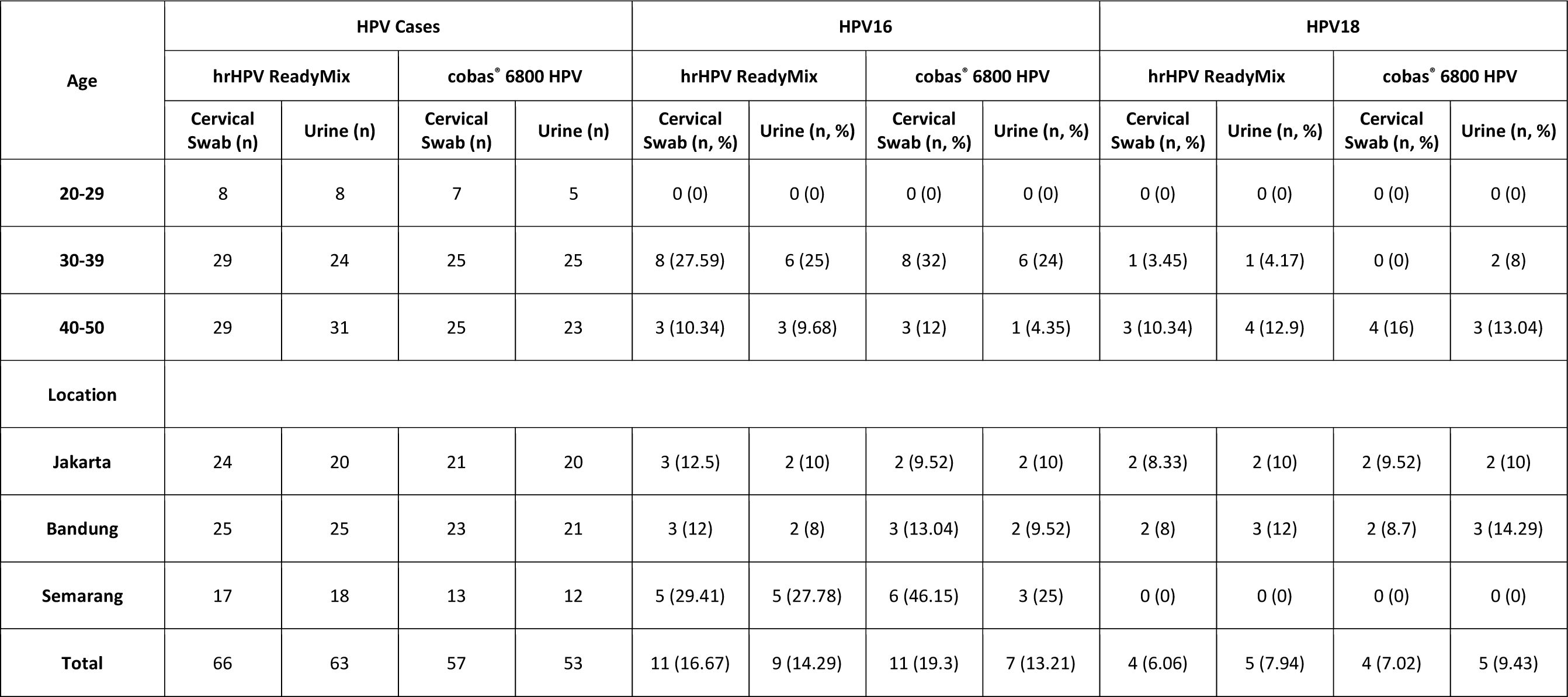
HPV16 and HPV18 proportion within all HPV cases.

## Notes

### Author Declarations

Health Research Ethics Committee of the Faculty of Medicine Universitas Indonesia Dr. Cipto Mangunkusumo Hospital gave ethical approval of this work under the identifier No. KET- 674 / UN2.F1/ETIK/PPM.00.02/2022.

### Summary of Updates

Updated title on the manuscript, grammatical error correction, and references update

