## Supplementary Data for "Urinary high-risk HPV DNA detection to enhance cervical cancer screening in developing countries"

Of

| A) | 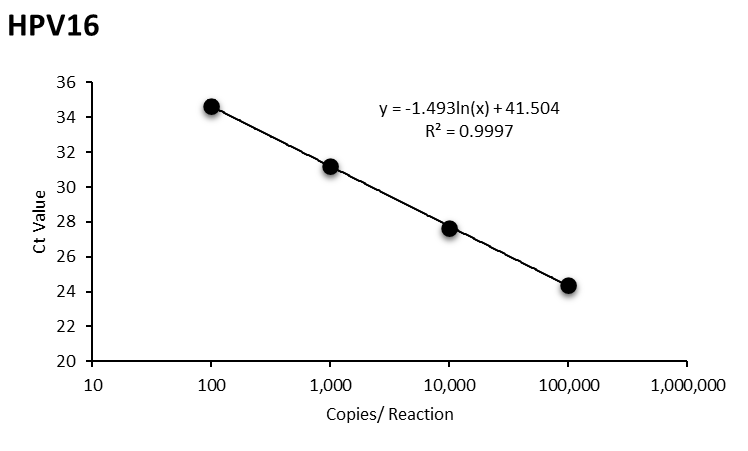 | H) | 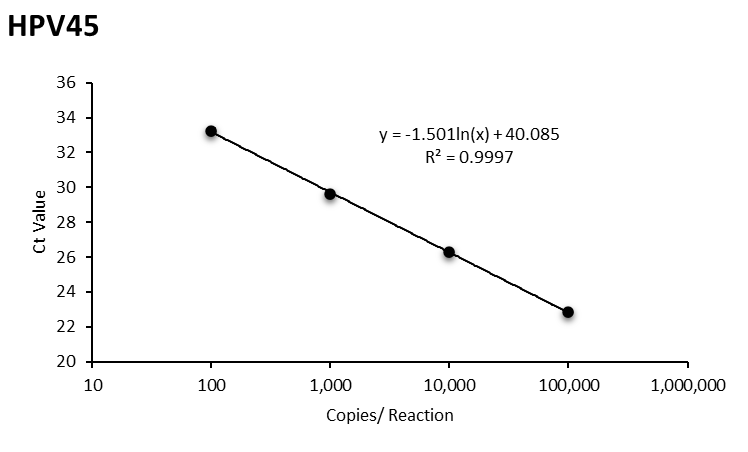 |
| --- | --- | --- | --- |
| B) | 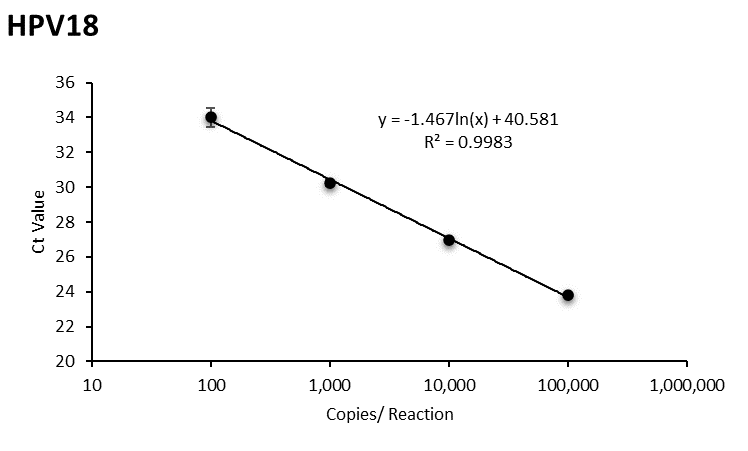 | I) | 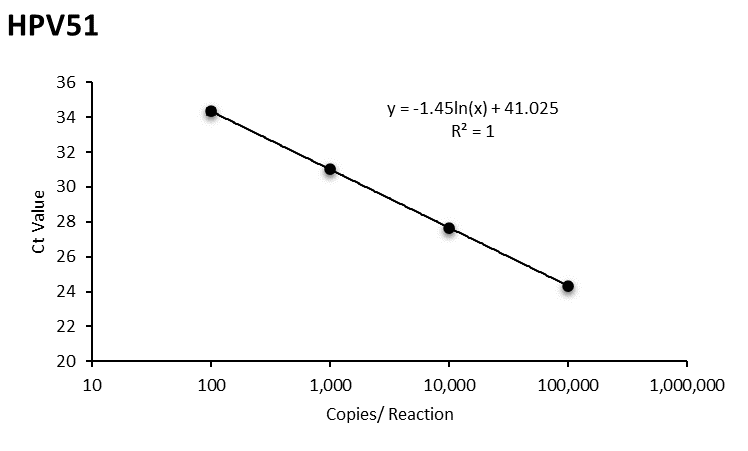 |
| C) | 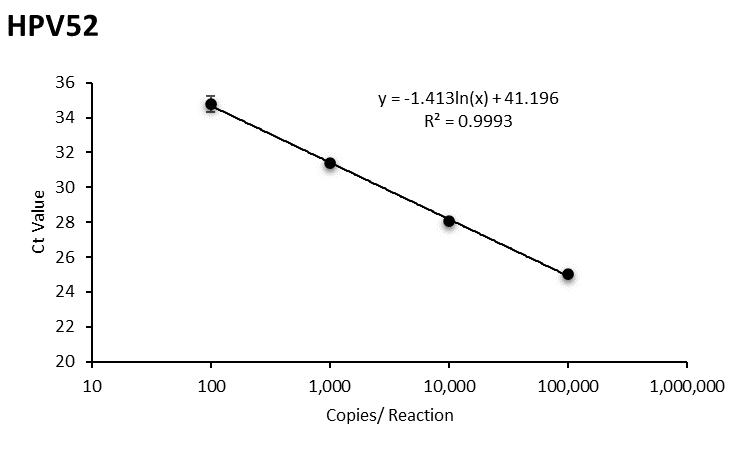 | J) | 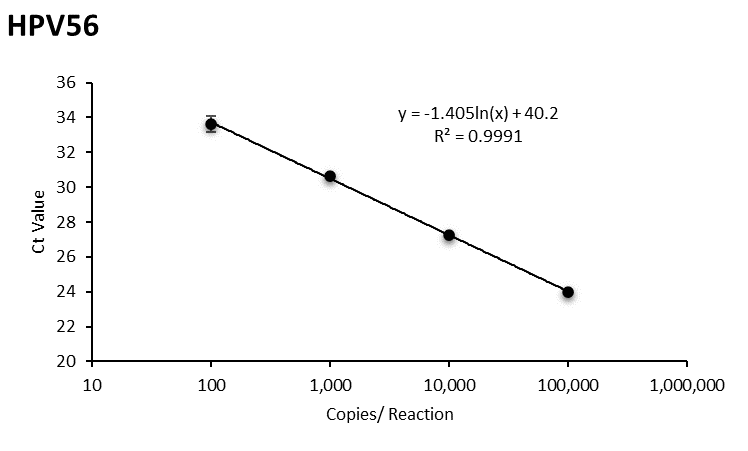 |
| D) | 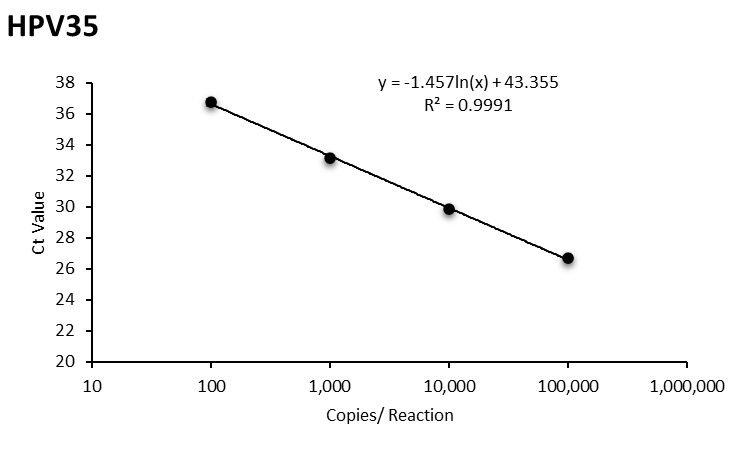 | K) | 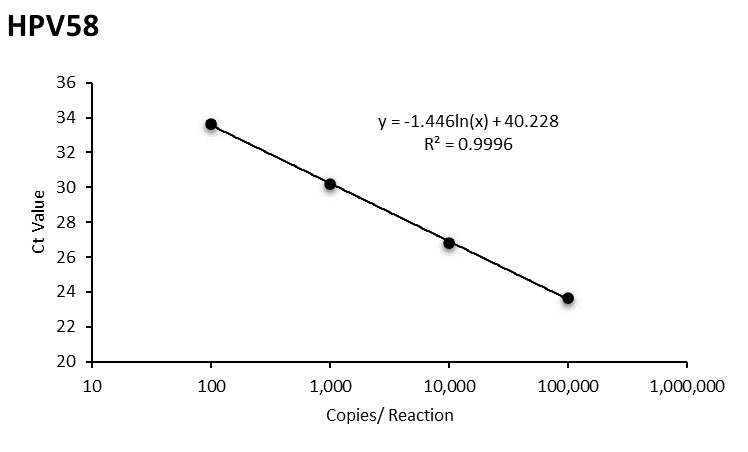 |
| E) | 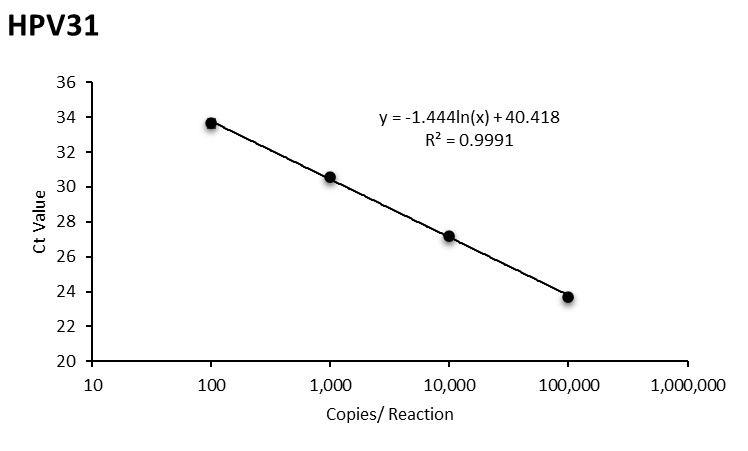 | L) | 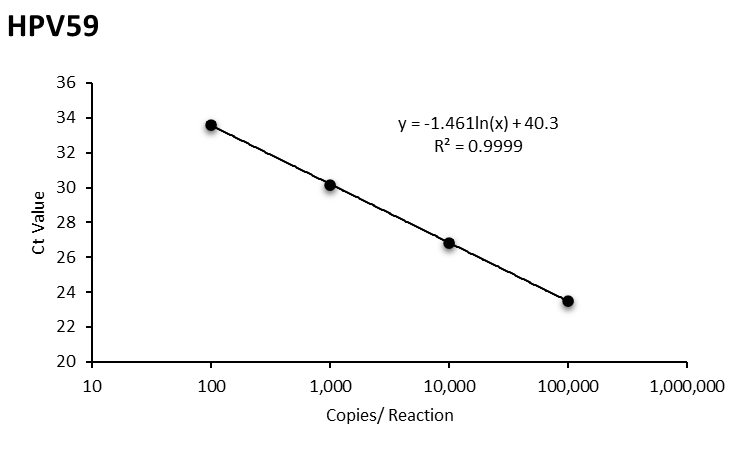 |
| F) | 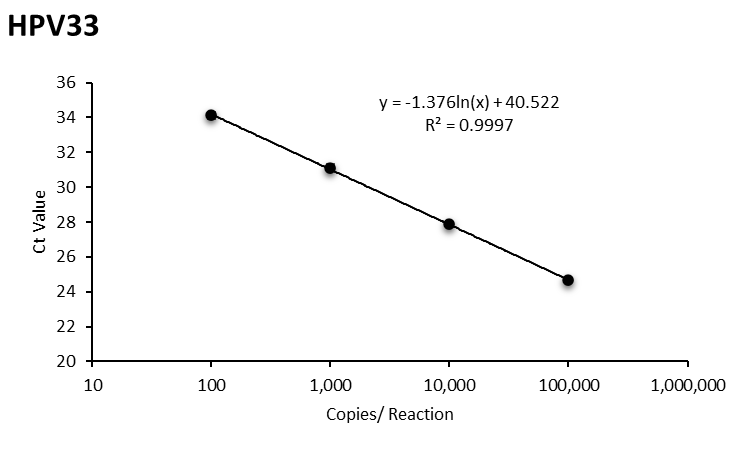 | M) | 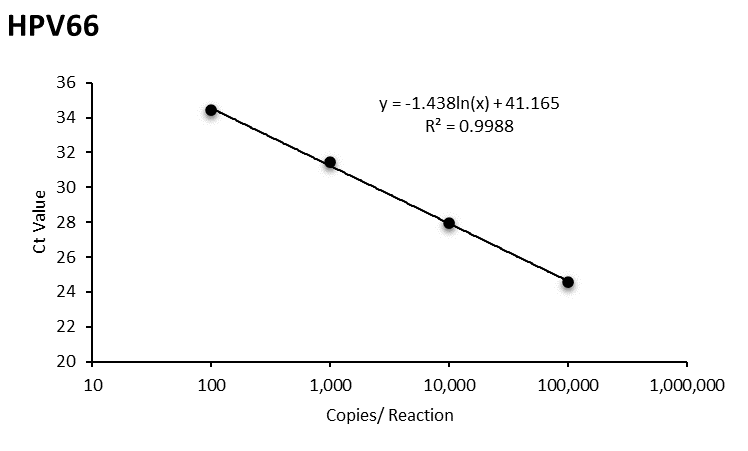 |
| G) | 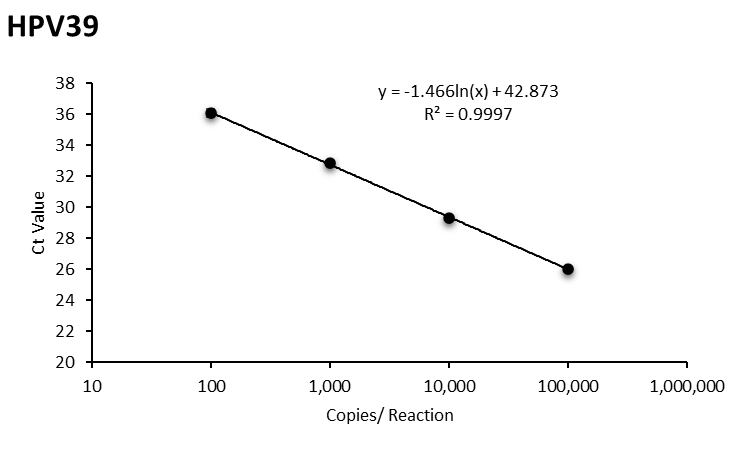 | N) | 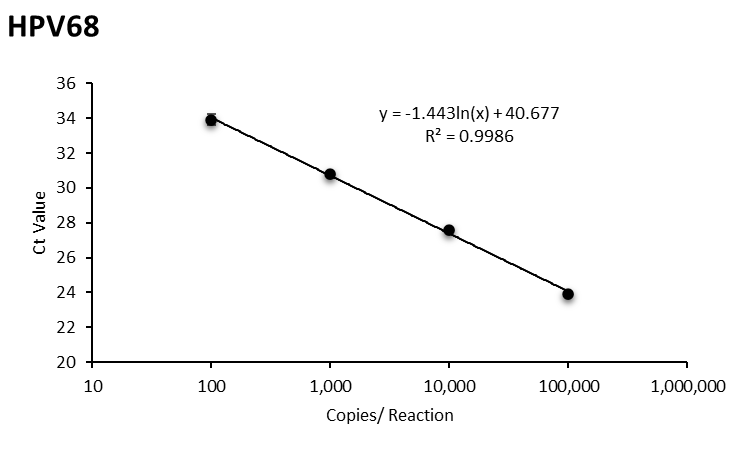 |

Supplementary Fig S1. Linearity assay of ReadyMix in detecting 14 hrHPV types using HPV synthetic DNA as template ranging from 100 to 100,000 GE copies/reaction. A) HPV16; B) HPV18; C) HPV52; D) HPV35; E) HPV31; F) HPV33; G) HPV39; H) HPV45; I) HPV51; J) HPV56; K) HPV58; L) HPV59; M) HPV66; N) HPV68. This assay demonstrated excellent efficiency with a range between 90-110% and R^2^ ~1.

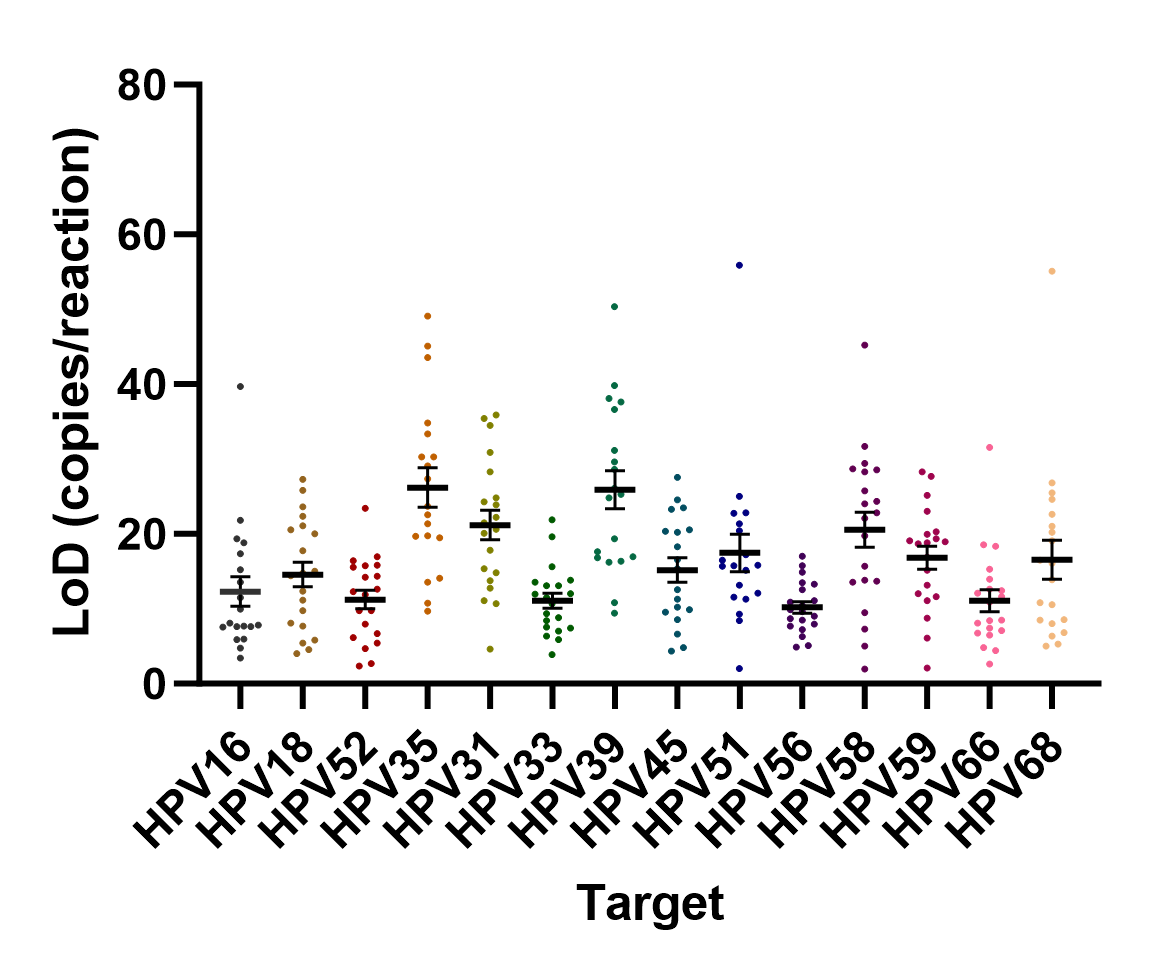

Supplementary Fig S2. Limit of Detection (LoD) of ReadyMix for each hrHPV target ranging from 10-28 copies/20 uL qPCR reaction

Supplementary Table S1. The results of NGS run to investigate the presence of hrHPV in the samples based on E6-E7 amplicon sequencing

| No | Seq ID | Sample Type | False Positive to | | | qPCR Result | hrHPV Detected (%) |
| --- | --- | --- | --- | --- | --- | --- | --- |
|  |  |  | Cervical Swab cobas | Cervical Swab ReadyMix | Urine  cobas |  |  |
| 1 | 140-A-E | Cervical Swab | V | - | - | HPV Other | HPV68 (81.1%), HPV18 (5.7%), HPV51 (4.9%), HPV45 (1.8%), HPV16 (1.8%), HPV82 (1.3%), HPV52 (0.9%), HPV33 (0.5%) |
| 2 | 313-A-E | Cervical Swab | V | - | - | HPV16 | HPV16 (59.3%), HPV52 (20.1%), HPV51 (6.7%), HPV33 (6.1%), HPV31 (3.7%), HPV18 (1.7%), HPV56 (1.3%) |
| 3 | 367-A-E | Cervical Swab | V | - | - | HPV52 | HPV52 (98.5%) |
| 4 | 645-A-E | Cervical Swab | V | - | - | HPV Other | HPV33 (97.6%), HPV16 (1.2%) |
| 5 | 811-A-E | Cervical Swab | V | - | - | HPV Other | HPV56 (57.5%), HPV51 (41.0%) |
| 6 | 904-A-E | Cervical Swab | V | - | - | HPV Other | HPV51 (94.6%), HPV56 (1.6%), HPV52 (1.3%), HPV33 (1.1%) |
| 7 | 140-U-E | Urine | V | - | - | HPV Other | HPV68 (98.6%) |
| 8 | 232-U-E | Urine | - | - | V | HPV Other | HPV59 (74.2%), HPV16 (7.1%), HPV51 (4.2%), HPV58 (3.8%), HPV68 (2.7%), HPV18 (1.5%), HPV31 (1.2%), HPV35 (1.2%), HPV56 (1.1%), HPV45 (0.9%), HPV52 (0.8%) |
| 9 | 313-U-E | Urine | V | - | V | HPV16 | HPV16 (90.6%), HPV58 (2.8%), HPV68 (1.9%), HPV59 (1.9%), HPV33 (1.4%) |
| 10 | 367-U-E | Urine | V | - | - | HPV52, Other | HPV52 (55.7%), HPV56 (44.1%) |
| 11 | 548-U-E | Urine | V | V | - | HPV 52 | HPV52 (91.2%), HPV51 (4.1%), HPV16 (1.7%), HPV18 (1.4%) |
| 12 | 043-U-E | Urine | - | - | - | HPV 18 | HPV18 (93.8%), HPV58 (2.2%), HPV33 (1.9%), HPV31 (1.0%) |
| 13 | 402-U-E | Urine | - | - | - | HPV16 | HPV16 (78.1%), HPV39 (8.0%), HPV52 (4.5%), HPV51 (3.6%), HPV58 (2.7%), HPV33 (1.3%) |
| 14 | 410-U-E | Urine | - | - | - | HPV52 | HPV52 (99.3%) |
| 15 | 413-U-E | Urine | V | V | - | HPV52 | HPV52 (97.1%), HPV18 (0.7%), HPV31 (0.7%) |
| 16 | 191-U-E | Urine | V | V | - | HPV Other | HPV31 (95.0%), HPV51 (1.3%), HPV16 (1.0%), HPV35 (0.5%), HPV56 (0.4%) |
| 17 | 446-U-E | Urine | V | - | - | HPV52 | HPV52 (97.9%), HPV18 (1.0%) |
| 18 | 465-U-E | Urine | - | - | - | HPV52, Other | HPV52 (85.0%), HPV31 (14.1%) |
| 19 | 633-U-E | Urine | - | - | V | HPV Other | HPV51 (97.7%), HPV16 (1.0%) |
| 20 | 645-U-E | Urine | V | - | - | HPV Other | HPV33 (97.9%), HPV51 (0.8%) |
| 21 | 669-U-E | Urine | V | V | - | HPV 18 | HPV18 (94.0%), HPV31 (2.5%), HPV52 (0.9%), HPV16 (0.7%) |
| 22 | 673-U-E | Urine | V | V | - | HPV Other | HPV68 (95.2%), HPV16 (1.9%), HPV18 (1.4%) |
| 23 | 695-U-E | Urine | V | V | V | HPV Other | HPV33 (95.2%), HPV31 (1.9%), HPV52 (0.7%), HPV45 (0.6%) |
| 24 | 710-U-E | Urine | V | V | V | HPV Other | HPV39 (96.2%), HPV52 (1.5%), HPV33 (0.9%) |
| 25 | 718-U-E | Urine | - | V | - | HPV Other | HPV58 (96.7%), HPV51 (1.8%) |
| 26 | 787-U-E | Urine | - | - | V | HPV16, HPV Other | HPV58 (97.5%), HPV16 (1.0%) |
| 27 | 811-U-E | Urine | V | - | - | HPV Other | HPV56 (95.0%), HPV18 (2.1%), HPV51 (1.1%) |
| 28 | 816-U-E | Urine | - | - | - | HPV Other | HPV51 (97.6%), HPV52 (0.6%) |
| 29 | 825-U-E | Urine | V | V | V | HPV52 | HPV52 (98.0%) |
| 30 | 833-U-E | Urine | - | - | - | HPV16 | HPV16 (66.5%), HPV58 (8.5%), HPV18 (6.4%), HPV33 (4.3%), HPV52 (3.2%), HPV45 (3.2%), HPV51 (2.7%), HPV56 (2.7%), HPV68 (2.1%) |
| 31 | 889-U-E | Urine | V | V | - | HPV Other | HPV45 (89.3%), HPV56 (3.6%), HPV51 (3.0%), HPV52 (1.1%), HPV33 (0.5%), HPV18 (0.5%) |
| 32 | 305-A-E | Cervical Swab | N/A | | | HPV16 | HPV16 (52.8%), HPV18 (32.6%), HPV33 (4.9%), HPV51 (3.1%), HPV31 (1.4%), HPV59 (1.4%), HPV56 (1.0%), HPV39 (1.0%) |
| 33 | 616-A-E | Cervical Swab |  |  |  | HPV Other | HPV68 (83.5%), HPV16 (6.9%), HPV52 (3.0%), HPV51 (2.3%), HPV56 (2.1%), HPV82 (1.3%) |
| 34 | 253-A-E | Cervical Swab |  |  |  | HPV52, Other | HPV52 (85.3%), HPV51 (13.2%) |
| 35 | 189-A-E | Cervical Swab |  |  |  | HPV52 | HPV52 (64.7%), HPV82 (33.9%) |
| 36 | 465-A-E | Cervical Swab |  |  |  | HPV Other | HPV31 (91.7%), HPV51 (1.6%), HPV33 (1.4%), HPV56 (1.2%), HPV18 (1.0%), HPV35 (0.6%), HPV52 (0.5%) |
| 37 | 471-A-E | Cervical Swab |  |  |  | HPV52, HPV Other | HPV52 (55.4%), HPV56 (28.4%), HPV51 (14.9%) |
| 38 | 498-A-E | Cervical Swab |  |  |  | HPV18, HPV Other | HPV33 (58.5%), HPV18 (40.6%) |
| 39 | 823-U-E | Urine |  |  |  | HPV16, HPV Other | HPV51 (89.8%), HPV16 (8.5%) |
| 40 | 189-U-E | Urine |  |  |  | HPV52 | HPV52 (99.2%) |
| 41 | CA1-A-E | Cervical Swab |  |  |  | HPV Other | HPV59 (78.5%), HPV18 (4.2%), HPV52 (3.8%), HPV33 (2.8%), HPV82 (2.6%), HPV51 (1.8%), HPV31 (1.4%), HPV58 (1.4%), HPV16 (0.9%), HPV39 (0.9%) |
| 42 | CA1-U-E | Urine |  |  |  | HPV Other | HPV59 (91.2%), HPV33 (4.7%), HPV58 (2.3%) |
| 43 | gBlocks | gBlocks |  |  |  | HPV16, HPV18, HPV52, and HPV Other | HPV18 (20.6%), HPV35 (14.1%), HPV45 (13.3%), HPV58 (10.1%), HPV33 (9.6%), HPV56 (8.7%), HPV51 (7.9%), HPV59 (5.5%), HPV66 (4.2%), HPV52 (2.3%), HPV31 (2.3%), HPV68 (0.88%), HPV16 (0.30%), HPV39 (0.27%). |

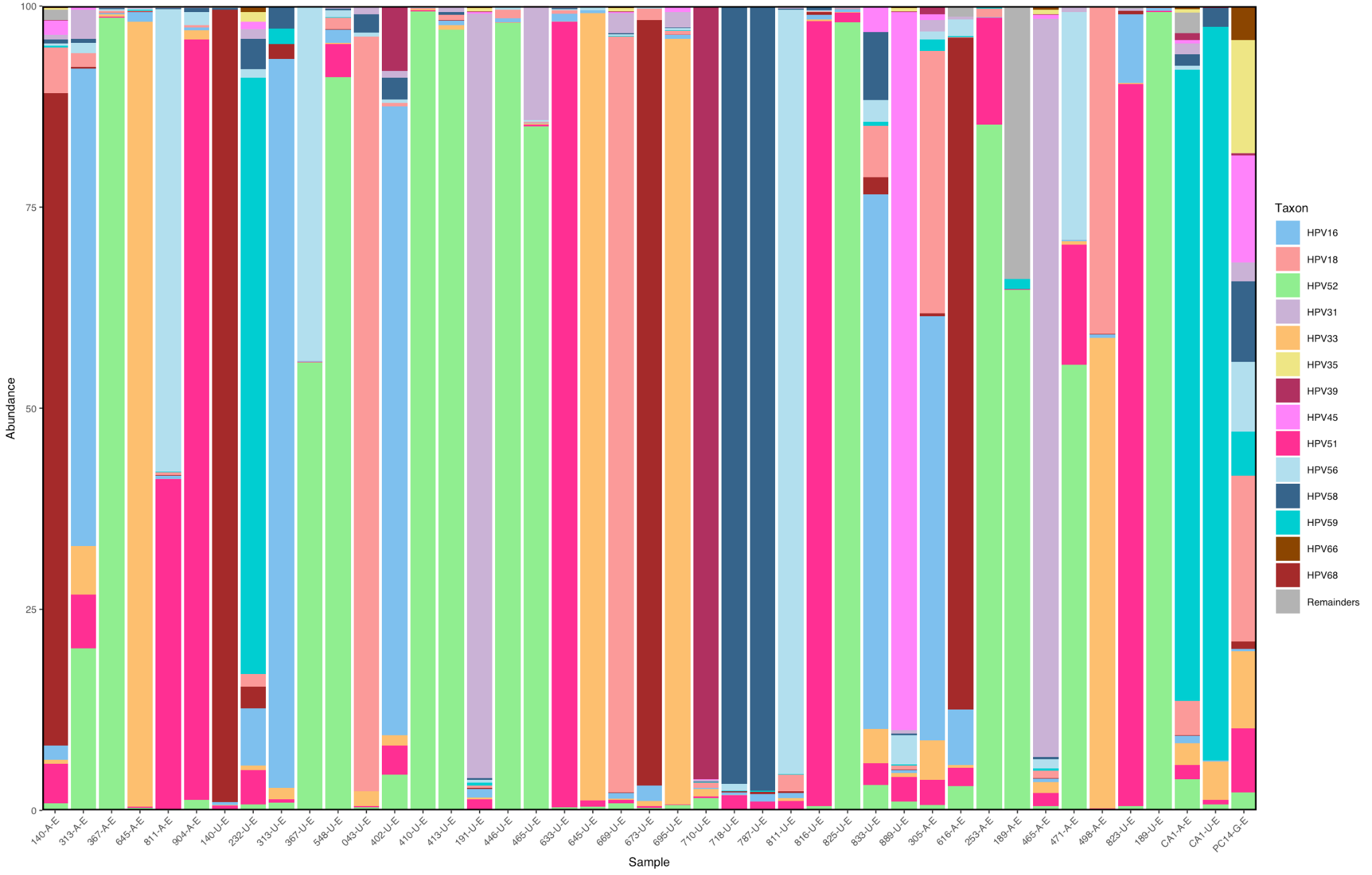

| Output | %>Q30 | Data with > Q30 | Total reads | Reads identified | Number of samples | Average reads/sample |
| --- | --- | --- | --- | --- | --- | --- |
| 0.66 Gb | 83.08% | 0.54 Gb | 1.704.265 | 217.015 | 42 | 5.047 |

Supplementary Fig S3. Stacked taxa bar plot showing the relative abundance of HPV type found in the sample

Supplementary Table S2. HPV detection using cobas in urine versus cervical swab and NGS

| **Urine cobas** | **Cervical Swab cobas & NGS** | | | **k-coefficient** |
| --- | --- | --- | --- | --- |
|  | **Positive** | **Negative** | **Total** |  |
| Positive | 47 | 2 | 49 | 0.84 |
| Negative | 14 | 588 | 602 |  |
| Total | 61 | 590 | 651 |  |
| **Effect size** | **Value (%)** | **95% CI (%)** | |  |
| Sensitivity | 77.05 | 65.09 - 85.81 | |  |
| Specificity | 99.66 | 98.77 - 99.94 | |  |
| Positive Predictive Value | 95.92 | 86.29 - 99.27 | |  |
| Negative Predictive Value | 97.67 | 96.13 - 98.61 | |  |
| Accuracy | 97.54 | 96.04 - 98.48 | |  |

Supplementary Table S3. Demographics of the study population

| **Age Range**  **Average Age** | **(mean** ± SD) | **Total (n)** | **Proportion (%)** | **Prevalence of HPV Type/ Group** | | | | | | | | | | | | | | | | | |
| --- | --- | --- | --- | --- | --- | --- | --- | --- | --- | --- | --- | --- | --- | --- | --- | --- | --- | --- | --- | --- | --- |
|  |  |  |  | **hrHPV ReadyMix** | | | | | | | | | | **cobas^®^ 6800 HPV** | | | | | | | |
|  |  |  |  | **Cervical Swab (n, %)** | | | | | **Urine (n, %)** | | | | | **Cervical Swab (n, %)** | | | | **Urine (n, %)** | | | |
|  |  |  |  | **HPV16** | **HPV18** | **HPV52** | **HPV Others** | **Coinfection** | **HPV16** | **HPV18** | **HPV52** | **HPV Others** | **Coinfection** | **HPV16** | **HPV18** | **HPV Others** | **Coinfection** | **HPV16** | **HPV18** | **HPV Others** | **Coinfection** |
| **20-29** | 26.45 ± 2.46 | 157 | 17.92% | 0 (0) | 0 (0) | 1 (0.64) | 3 (1.91) | 2 (1.27) | 0 (0) | 0 (0) | 2 (1.27) | 2 (1.27) | 2 (1.27) | 0 (0) | 0 (0) | 7 (4.46) | 0 (0) | 0 (0) | 0 (0) | 5 (3.18) | 0 (0) |
| **30-39** | 34.59 ± 2.99 | 346 | 39.50% | 7 (2.02) | 0 (0) | 6 (1.73) | 9 (2.6) | 3 (0.87) | 4 (1.16) | 1 (0.29) | 4 (1.16) | 9 (2.6) | 3 (0.87) | 6 (1.73) | 0 (0) | 15 (4.34) | 2 (0.58) | 5 (1.45) | 2 (0.58) | 16 (4.62) | 1 (0.29) |
| **40-50** | 44.23 ± 3.00 | 373 | 42.58% | 2 (0.54) | 3 (0.8) | 3 (0.8) | 17 (4.56) | 2 (0.54) | 2 (0.54) | 3 (0.8) | 6 (1.61) | 14 (3.75) | 3 (0.8) | 1 (0.27) | 3 (0.8) | 15 (4.02) | 3 (0.8) | 1 (0.27) | 2 (0.54) | 18 (4.83) | 1 (0.27) |
| **Location** |  | | | | | | | | | | | | | | | | | | | | |
| **Jakarta** | N/A | 385 | 43.95% | 3 (0.78) | 2 (0.52) | 6 (1.56) | 11 (2.86) | 1 (0.26) | 2 (0.52) | 2 (0.52) | 7 (1.82) | 9 (2.34) | 0 (0) | 2 (0.52) | 2 (0.52) | 17 (4.42) | 0 (0) | 2 (0.52) | 2 (0.52) | 16 (4.16) | 0 (0) |
| **Bandung** | N/A | 355 | 40.53% | 3 (0.85) | 1 (0.28) | 3 (0.85) | 14 (3.94) | 2 (0.56) | 2 (0.56) | 2 (0.56) | 3 (0.85) | 12 (3.38) | 3 (0.85) | 2 (0.56) | 1 (0.28) | 16 (4.51) | 2 (0.56) | 2 (0.56) | 2 (0.56) | 15 (4.23) | 1 (0.28) |
| **Semarang** | N/A | 136 | 15.53% | 3 (2.21) | 0 (0) | 1 (0.74) | 4 (2.94) | 4 (2.94) | 2 (1.47) | 0 (0) | 2 (1.47) | 4 (2.94) | 5 (3.68) | 3 (2.21) | 0 (0) | 4 (2.94) | 3 (2.21) | 2 (1.47) | 0 (0) | 8 (5.88) | 1 (0.74) |
| **Total** | 37.23 ± 7.28 | 876 | 100.00% | 9 (1.03) | 3 (0.34) | 10 (1.14) | 29 (3.31) | 7 (0.8) | 6 (0.68) | 4 (0.46) | 12 (1.37) | 25 (2.85) | 8 (0.91) | 7 (0.8) | 3 (0.34) | 37 (4.22) | 5 (0.57) | 6 (0.68) | 4 (0.46) | 39 (4.45) | 2 (0.23) |

Supplementary Table S4. HPV16 and HPV18 proportion within all HPV cases

| **Age** | **HPV Cases** | | | | **HPV16** | | | | **HPV18** | | | |
| --- | --- | --- | --- | --- | --- | --- | --- | --- | --- | --- | --- | --- |
|  | **hrHPV ReadyMix** | | **cobas^®^ 6800 HPV** | | **hrHPV ReadyMix** | | **cobas^®^ 6800 HPV** | | **hrHPV ReadyMix** | | **cobas^®^ 6800 HPV** | |
|  | **Cervical Swab (n)** | **Urine (n)** | **Cervical Swab (n)** | **Urine (n)** | **Cervical Swab (n, %)** | **Urine (n, %)** | **Cervical Swab (n, %)** | **Urine (n, %)** | **Cervical Swab (n, %)** | **Urine (n, %)** | **Cervical Swab (n, %)** | **Urine (n, %)** |
| **20-29** | 8 | 8 | 7 | 5 | 0 (0) | 0 (0) | 0 (0) | 0 (0) | 0 (0) | 0 (0) | 0 (0) | 0 (0) |
| **30-39** | 29 | 24 | 25 | 25 | 8 (27.59) | 6 (25) | 8 (32) | 6 (24) | 1 (3.45) | 1 (4.17) | 0 (0) | 2 (8) |
| **40-50** | 29 | 31 | 25 | 23 | 3 (10.34) | 3 (9.68) | 3 (12) | 1 (4.35) | 3 (10.34) | 4 (12.9) | 4 (16) | 3 (13.04) |
| **Location** |  | | | | | | | | | | | |
| **Jakarta** | 24 | 20 | 21 | 20 | 3 (12.5) | 2 (10) | 2 (9.52) | 2 (10) | 2 (8.33) | 2 (10) | 2 (9.52) | 2 (10) |
| **Bandung** | 25 | 25 | 23 | 21 | 3 (12) | 2 (8) | 3 (13.04) | 2 (9.52) | 2 (8) | 3 (12) | 2 (8.7) | 3 (14.29) |
| **Semarang** | 17 | 18 | 13 | 12 | 5 (29.41) | 5 (27.78) | 6 (46.15) | 3 (25) | 0 (0) | 0 (0) | 0 (0) | 0 (0) |
| **Total** | 66 | 63 | 57 | 53 | 11 (16.67) | 9 (14.29) | 11 (19.3) | 7 (13.21) | 4 (6.06) | 5 (7.94) | 4 (7.02) | 5 (9.43) |
